# Effectiveness and efficiency of immunisation strategies to prevent RSV among infants and older adults in Germany: a modelling study

**DOI:** 10.1101/2024.06.20.24309248

**Authors:** Fabienne Krauer, Felix Guenther, Marina Treskova-Schwarzbach, Viktoria Schoenfeld, Mihaly Koltai, Mark Jit, David Hodgson, Udo Schneider, Ole Wichmann, Thomas Harder, Frank G. Sandmann, Stefan Flasche

## Abstract

**Background:** Recently, several novel RSV immunisation products that protect infants and older adults against RSV disease have been licensed in Europe. We estimated the effectiveness and efficiency of introducing these RSV immunisation strategies in Germany.

**Methods:** We used a Bayesian framework to fit an age-structured dynamic transmission model of RSV to sentinel surveillance and RSV-specific hospitalisation data in Germany from 2015-2019. The calibrated model was used to evaluate different RSV intervention strategies over five years: long-acting, single-dose monoclonal antibodies (mAbs) in high-risk infants aged 1-5 months; long-acting mAbs in all infants aged 1-5 months; seasonal vaccination of pregnant women and one-time seasonal vaccination of older adults (75+/65+/55+ years). We performed sensitivity analysis on vaccine uptake, seasonal vs. year-round maternal vaccination, and the effect of under-ascertainment for older adults.

**Results:** The model was able to match the various RSV datasets. Replacing the current short-acting mAB for high-risk infants with long-acting mAbs prevented 1.1% of RSV-specific hospitalisations in infants per year at the same uptake. Expanding the long-acting mAB programme to all infants prevented 39.3% of infant hospitalisations per year. Maternal vaccination required a larger number to be immunised to prevent one additional hospitalisation than a long-acting mAB for the same uptake. Vaccination of adults older than 75 years at an uptake of 40% in addition to Nirsevimab in all infants prevented an additional 4.5% of all RSV-hospitalisations over five years, with substantial uncertainty in the correction for under-ascertainment of the RSV burden.

**Conclusions:** Immunisation has the potential to reduce the RSV disease burden in Germany.

## Introduction

Respiratory syncytial virus (RSV) infection is a major health problem globally and can cause severe disease in infants and older adults. Annually, an estimated 3.6 million children globally are admitted to hospital for RSV-associated lower respiratory tract infection and over 100,000 die ^1^. In high-income countries, almost half a million adults aged 60 years or older are estimated to be hospitalised each year due to RSV ^2^. In Germany, the annual hospitalisation rate in <2 year-olds is at least 28.6 per 1000 population ^3^ (almost 46,000 infants per year). For older adults, the estimated hospitalisations are ~5,400 per year in 65-74-year-old and ~14,000 in 75 years and older ^4^, although these numbers may underestimate the true disease burden due to underdiagnosing of RSV in older adults.

For infants, the current strategy for RSV prevention relies on the monoclonal antibody Palivizumab, which is administered as a series of up to five injections in one-month intervals during RSV seasons. In Germany, Palivizumab has been indicated since 2002 for use in young infants with high risk of severe disease (born before gestational week 35, and younger than 6 months at the beginning of the season, and infants up to 2 years born with bronchopulmonary dysplasia (BPD) or congenial heart disease (CHD)) ^5^. Until recently, no other product was available to protect infants or adults from RSV.

In 2023, a novel long-lasting monoclonal antibody, Nirsevimab (Beyfortus®), as well as a maternal vaccine, Abrysvo®, have been licensed in the EU for the prevention of RSV. Nirsevimab is given as a single dose in early infancy and has been shown to prevent about 70-80% of RSV-associated hospital admissions in healthy young children, including those born prematurely, for the first 150 days after immunisation ^6–8^. Abrysvo® is administed during pregnancy and was found to have a 70% efficacy in preventing severe medically attended RSV-associated lower respiratory tract diseases in term born infants in the first 180 days of life ^9^. Abrysvo® is also licensed to immunise older adults, together with another vaccine, Arexvy®. Both showed vaccine efficacy (VE) of at least 66% against lower respiratory tract infection (LRTI) ^10,11^, and an continued effect into the second season after administration ^12,13^.

In this study, we used mathematical modelling to synthesize evidence on the RSV-associated burden of disease in Germany and used this framework to estimate the potential impact of different RSV immunisation strategies, which use these newly available products to prevent RSV disease in young children and older adults.

## Methods

### RSV burden estimates

#### Sentinel surveillance data from primary care providers

We used data from the ‘Arbeitsgemeinschaft Influenza’ (AGI), which is a sentinel network for acute respiratory infections (ARI) in a primary care setting. This sentinel network of around 700 paediatricians and general practitioners covers about 1% of the population of Germany. Participating physicians report the number of cases with ARI. A proportion of these cases are swabbed (nasal or nasopharyngeal) and tested through PCR for various respiratory viruses including RSV. For this study, we used only the weekly number of laboratory-confirmed RSV cases, which were originally stratified by broad age groups (<1 year, 1-2 years, 2-4 years, 5-14 years, 15-34 years, 35-49 years, 50-59 years, 60+ years). Due to the small numbers in the individual age groups, we fit the model to the total of the weekly cases as well as to the proportional distribution of the different age groups (over all time points).

#### Hospitalisation data

We also used RSV-specific hospitalisation data from the largest health insurance company in Germany, the ‘Techniker Krankenkasse’ (TK). The data comprise all insurance claims submitted to TK between 2015–2019 in an inpatient setting and with an RSV-specific ICD10 code as the main or secondary diagnosis. The selected RSV-specific ICD-10 codes were J12.1, 20.5, J21.0, B97.4. The data were stratified by 25 age groups to match the age structure in the model (see model structure). The hospitalisation case data were extrapolated to the total population of Germany adjusting for the different coverages by TK in the different age groups. For the fitting, the final dataset was aggregated twofold. First, we summed the hospitalizations over all age groups and fitted it by quarter and year, and second, we summed the hospitalisations over time and fitted the proportional distribution of the age group.

#### Intensive care unit (ICU) and in-hospital mortality data

To quantify ICU stays and in-hospital mortality, we sourced routinely collected hospitalisation data from the ‘Institut fuer das Entgeltsystem im Krankenhaus’ (InEK). We used primary diagnosis codes of yearly RSV-specific admissions to all hospitals in Germany in 2019-2023 to estimate age-specific hospitalisation-ICU and hospitalisation-mortality ratios. These are used as scaling factors for a more refined quantification of disease burden from our dynamic transmission model (for details see supplement).

#### Seroconversion data

For a better calibration of the model in the infant age groups, we additionally used seroconversion data from a cross-sectional seroprevalence study in the Netherlands (named “Pienter”). ^14^ These data were aggregated in monthly age groups for months 0-11 and the proportion of not-yet-seroconverted individuals was calculated.

#### Under-ascertainment of RSV hospitalisations in older adults

RSV is strongly underdiagnosed in older adults ^15^. We therefore assumed that the RSV-specific hospitalisations only represent a small fraction of the true burden. To account for under-ascertainment, we multiplied the RSV-specific hospitalisations in older adults simulated in the vaccination strategies with a scaling factor to estimate the true burden (for details see supplement). The scaling factor was estimated as the ratio of age-specific published RSV-related hospitalisations and our age-specific model-derived hospitalisations.

#### Model structure

We defined a deterministic compartmental ODE model stratified by age, number of past RSV infections and immunisation status (Fig. 1). For the full list of equations, see supplementary tables S2-S8. The 25 age groups include monthly age groups for up to one year, yearly age groups for up to 5 years, 5-year intervals for up to 15 years and 10-year intervals for 15+ year-old. The epidemiological states in the model represent maternal immunity after birth (M), susceptibility (S), latent infection (E), symptomatic infectiousness (I), asymptomatic infectiousness (A) and temporary but full immunity (R). To allow for realistic durations of immunity, we split the maternal and immune/recovered compartments (M1, M2, R1, R2) and divided the corresponding rates by 2 to allow for an Erlang distributed duration of immunity. Susceptible individuals become infected (S −> E) and symptomatically (E −> I) or asymptomatically (E −> A) infectious, and recover with temporary but full immunity (I or A −> R1 −> R2). After waning of immunity, they can become susceptible again (R2 −> S). Infants are born with maternal protection from infection (M1) if the mother was immune during the third trimester, otherwise infants are born unprotected (S). Maternal immunity (M1, M2) is considered full protection against infection during a short time period. The model also comprises 6 mutually exclusive immunisation status arms: non-immunised/unvaccinated individuals (1), infants immunised with mAbs (passive immunitsation) (2), vaccinated individuals (active immunisation) (3), infants immunised through maternal vaccination (passive immunisation) (4), unvaccinated pregnant women in the third trimester (5), vaccinated pregnant women in the third trimester (active immunisation) (6). The immunisation status arms also have a W compartment to allow for waning of immunity after vaccination or mAbs. Individuals move between age groups, states, levels and arms at rates specified in supplementary tables S9-S12. For a detailed description of the structure, see supplement. All analyses were performed in the programming languages Julia and R. Simulated model outputs (such as numbers of hospitalisations) are summarised by median and 2.5^th^ and 97.5^th^ percentile of the prediction interval (PI), marginal posterior estimates are summarised as median and 95% credible intervals (CrI). All code is available in a public GitHub repository (https://github.com/fkrauer/RSV-VACC-DE).

**Figure 1.**
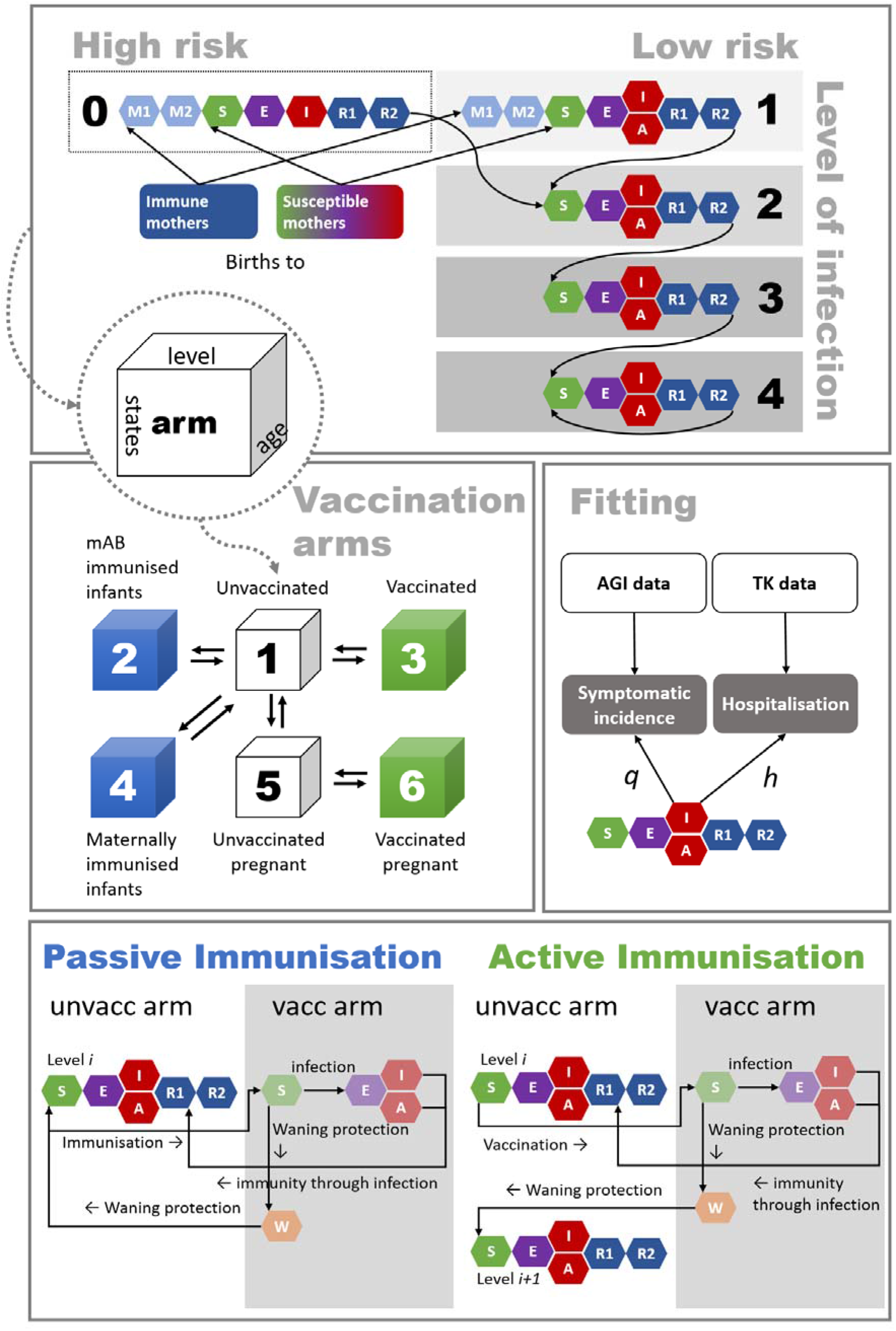
Model structure of RSV transmission and disease progression, and mechanisms of immunisation.

#### Model assumptions

- Individuals have 4 levels (1-4) of infection with decreasing susceptibility (i.e. increasing immune memory) after each re-infection. The number of levels of infection was chosen to match prior RSV models ^16,17^. Further reinfections are possible in the model but do not lead to additional immunity.
- A proportion of all infants are considered high risk (<=35 weeks gestational age, BPD, CHD) and are born into an extra level 0 (high risk, first infection), the rest are born into level 1 (low risk, first infection).
- High-risk infants have maternal immunity when born to mothers who were recently infected, but the duration of maternal protection is reduced by 50% compared to low-risk infants ^18^.
- High-risk infants also have a 3 times higher risk of hospitalisation in the first year of life than low-risk infants ^19–22^, and do not experience asymptomatic infection. After the first infection, high risk infants progress to the regular, second level of reinfection.
- The immunisation products do not prevent infection but reduce the risk of symptomatic disease and hospitalisation with efficacy estimates based on clinical trials.
- After the protection from an immunisation product wanes, individuals move back to the unvaccinated arms to either the same level of infection (for passive immunisation, i.e. no gain of immunity) or the next level of infection (for active immunisation, i.e. gain of immunity).
- Passive immunisation through maternal vaccination is assumed to initially lead to full protection against infection with the same duration as immunity from maternal infection, followed by a period of reduced probability of hospitalisation. In pre-term infants, maternal immunisation is assumed to have no additional effect besides the 50% reduced immunity of high risk infants.

#### Model fitting

The fitted model represents RSV epidemiology in Germany during the RSV seasons 2015-2019, when Palivizumab was given seasonally to high risk infants in months 1-5 of their life and with up to 5 monthly doses. For computational reasons, we assumed exponential waning of the protective effect of Palivizumab in the short interval between doses. The model was fitted to five datasets:

- Weekly incident outpatient cases, unstratified (“AGI”)
- Proportional distribution of the age groups among the incident outpatient cases (“AGI”)
- Quarterly hospitalisations, unstratified (“TK”)
- Proportional distribution of the age groups among the hospitalisations (“TK”)
- Proportion of not-yet-seroconverted (“Pienter”)

The weekly and quarterly time series data were fitted assuming a Negative-Binomial likelihood, the age distributions were fitted assuming a multinomial likelihood, the seroconversion data was fitted assuming a binomial likelihood. The total likelihood was calculated as the product of the individual likelihoods (see supplement for more details).

#### Immunisation strategies

In consultation with the working group of RSV at the German National Immunization Technical Advisory Group (“STIKO”), we modelled different RSV immunisation strategies (see Table 1). We considered two monoclonal antibodies (Palivizumab and Nirsevimab) for infants as well as a maternal and an elderly protein-based vaccine. We used the published estimates for VE and duration of protection for all immunisation products from clinical trials, and adjusted them for use in our model (see supplement text and table S12). The adjustment ensured that the average VE observed in the trial time period matched the average VE in the model resulting from an Erlang-2 distributed waning of protection. All strategies were modelled over a time horizon of five years. It was not possible to model a sequential roll-out of vaccination, instead we assumed that all eligible individuals could be immunised at the beginning of the season or as soon as they age into the eligible age group during the immunisation period.

**Table 1:** Overview of strategies of potential RSV immunisation strategies investigated for Germany.

| Strategy vs. comparator | Immunisation strategy | RSV intervention and eligible population | Assumed uptake |
| --- | --- | --- | --- |
| 0 (current) | Short-acting mAbs in high-risk infants | Palivizumab; high-risk infants aged 1-5 months; seasonal | 90% |
| 1 vs. 0 | Long-acting mAbs in high-risk infants | Nirsevimab, high-risk infants aged 1-5 months; seasonal | 90% |
| 2 vs. 0 | Long-acting mAbs in all infants | Nirsevimab, infants aged 1-5 months<sup>**</sup>; seasonal | 70%<sup>*</sup> |
| 3 vs. 0 | Maternal vaccination & Palivizumab in high risk infants | Vaccine, pregnant women aged 25-45 years; seasonal | 40%<sup>**</sup> |
| 4 vs. 2 | Older adult vaccination & Nirsevimab in all infants | Vaccine, single dose, adults aged 75+ years, administered during the season | 40%<sup>**</sup> |
| 5 vs. 2 | Older adult vaccination & Nirsevimab in all infants | Vaccine, single dose, adults aged 65+ years, administered during the season | 40%<sup>**</sup> |
| 6 vs. 2 | Older adult vaccination & Nirsevimab in all infants | Vaccine, single dose, adults aged 55+ years, administered during the season | 40%<sup>**</sup> |

<sup>\*</sup> Similar to Rotavirus/MenC vaccination in children in Germany.
<sup>\*\*</sup> Similar to Pertussis vaccination in pregnant women or Influenza vaccination in older adults in Germany.

### Infant RSV immunisation: Monoclonal antibodies and maternal vaccination

Infants are immunised seasonally from November to March with mAbs (both short and long-acting) in months 1-5 of their life (but not in the first 30 days of life) to reflect the current situation with Palivizumab where the median age at first injection is at 3.2 months (25^th^ percentile 1.8 months, 75^th^ percentile 5.5) ^23^. The maternal vaccine is modelled to be given seasonally from November to March to a proportion of pregnant people in the third trimester (25 to 45-year old women). The infant strategies were compared to the current base strategy (0).

### RSV vaccination of older adults

The vaccine in older adults is modelled to be given seasonally from October to February to individuals aged 75+, 65+, or 55+ year olds. Since there is currently no evidence of the effectiveness of booster doses, we modelled this strategy as a one-time single dose (rather than seasonally repeated like the influenza vaccine for example). In the first simulated year, all available individuals in the targeted age group are vaccinated simultaneously at the beginning of the year, after that only individuals ageing into the eligible age groups are vaccinated. RSV-specific hospitalisations in older adults were scaled up to account for under-ascertainment (see supplement). In addition to vaccinating older adults, we assumed that all infants aged month 1-5 were given Nirsevimab. The older adult strategies were thus compared to the infant strategy 2 (only Nirsevimab for all infants).

### Sensitivity analysis

We performed multiple additional sensitivity analyses to investigate the effect of central assumptions and certain specifications regarding the different immunisation strategies. First, we investigated the effect of varying the uptake for Nirsevimab in all infants and for the maternal vaccination. Second, we compared the effects of a seasonal maternal vaccination programme to a year-round strategy. For the older adult vaccination, we investigated different ages of eligibility and the under-ascertainment scaling factor, which we varied from 1 to 15.

## Results

### Model calibration

The model was able to fit the different datasets (Fig. 2): it replicated the seasonal pattern both in hospitalisations (Fig 2A) and outpatient numbers (Fig. 2C). Reported RSV hospitalisations at peak were lower in the first three seasons than in the last, which was not fully reflected in the model. The hospitalisation incidence in the last season however was matched well. Similarly, during 2016–19 there was an apparent biennial pattern in the number of outpatient RSV cases reported which was not replicated by the model (Fig. 2C), however the model matched the average seasonal pattern well. The symptomatic outpatient visits and the hospitalisation incidences by age group were not directly fitted (only indirectly through the proportion of cases in the individual age groups), but the model was still able to match these time series (Fig. S8). Model diagnostics confirmed that all chains converged (Fig. S9). The marginal posterior estimates are given in table S13.

**Figure 2.**
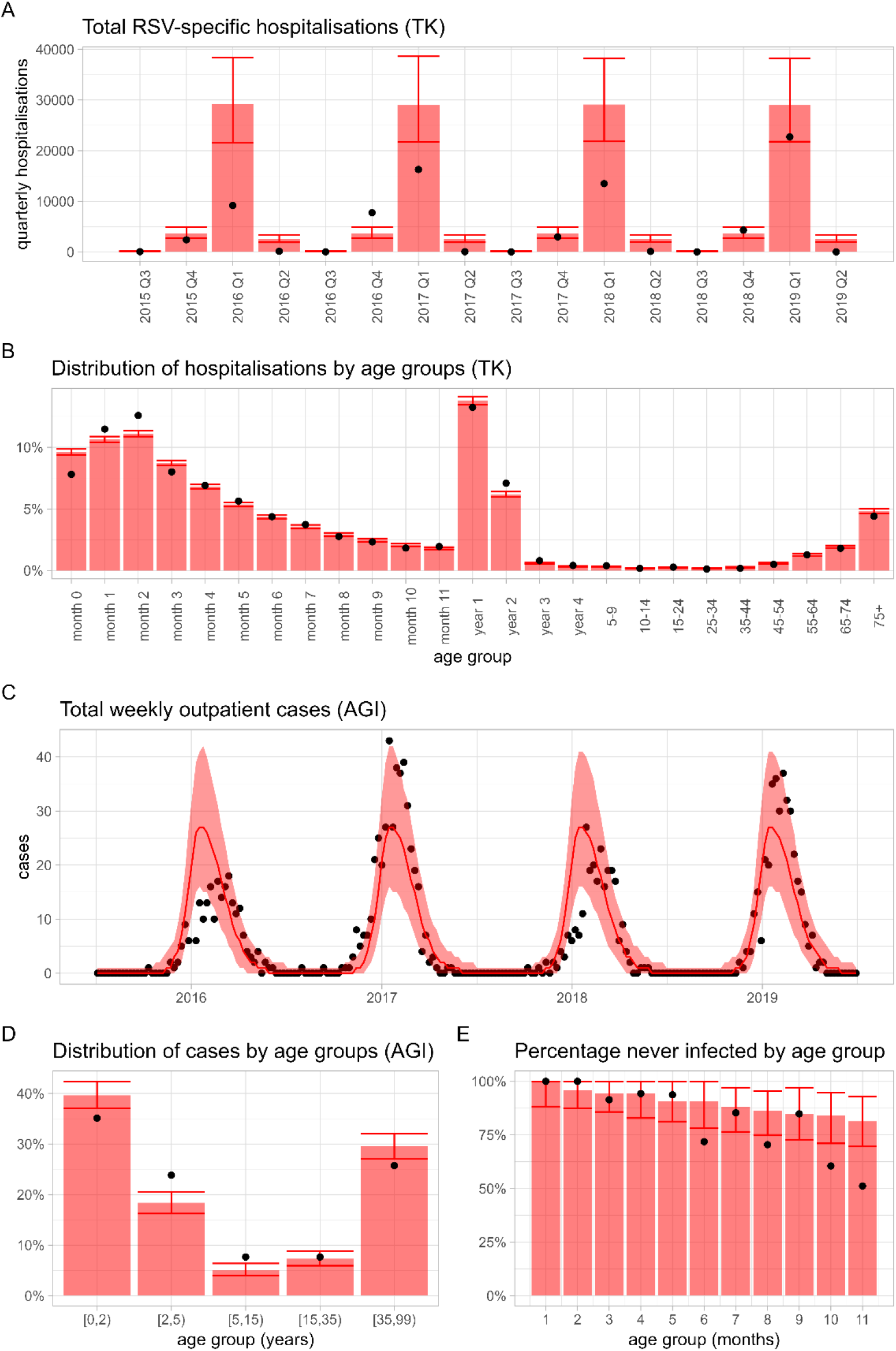
Fitting results of the dynamic transmission model to the data using Bayesian inference. The plots show the median model prediction and the 95% posterior prediction intervals (PPI) (red ribbon or error bar) compared to data (black points). The model was fitted to German RSV-specific hospitalisation data (TK, Fig. 2A, 2B), German sentinel outpatient RSV cases (AGI, Fig. 2C, 2D) and seroconversion data from the Netherlands in <1 year olds (2E).

### Current disease burden

We find that with the current RSV prevention strategy (Palivizumab for high risk infants), on average 12.5 million symptomatic cases, at-least 35,800 RSV-specific hospitalisations, 2600 intensive care (ICU) cases and 213 RSV-specific deaths occur in Germany each year in all age groups together (not accounting for under-ascertainment) (for age-specific estimates see supplementary table S14). 10.0-11.4 % of all hospitalisations in infants occur in high risk individuals. The probability of hospitalisation in high risk infants was estimated as 100% in the first three months of life but decreasing to 15% at month 11 (Fig. S10).

### Under-ascertainment in older adults

The comparison of published hospitalisation rates with our model derived estimates suggest that the RSV-specific ICD-10 codes underestimate the true burden in older adults of 55+ years by a factor 8 - 14 (Fig. S11). For older adults we thus scaled the hospitalisations and ICU admissions by a factor of 8 (strategies 1-3) or 8 and 14, respectively (strategies 4-6).

### Immunisation strategies to protect infants

Our results indicate that switching from Palivizumab to Nirsevimab for high-risk infants aged 1-5 months (strategy 1) would prevent only median 288 (95% PI 275-301) RSV-specific hospitalisations annually in all age groups (Figure 3A), corresponding to 0.79% of all RSV-specific hospitalisations (Fig. 3B) and 1.1% of all hospitalisations in <1 year old under the current strategy. This strategy prevents also 20 (95% PI 19-21) ICU cases (1.1% of infants in the ICU).

**Figure 3:**
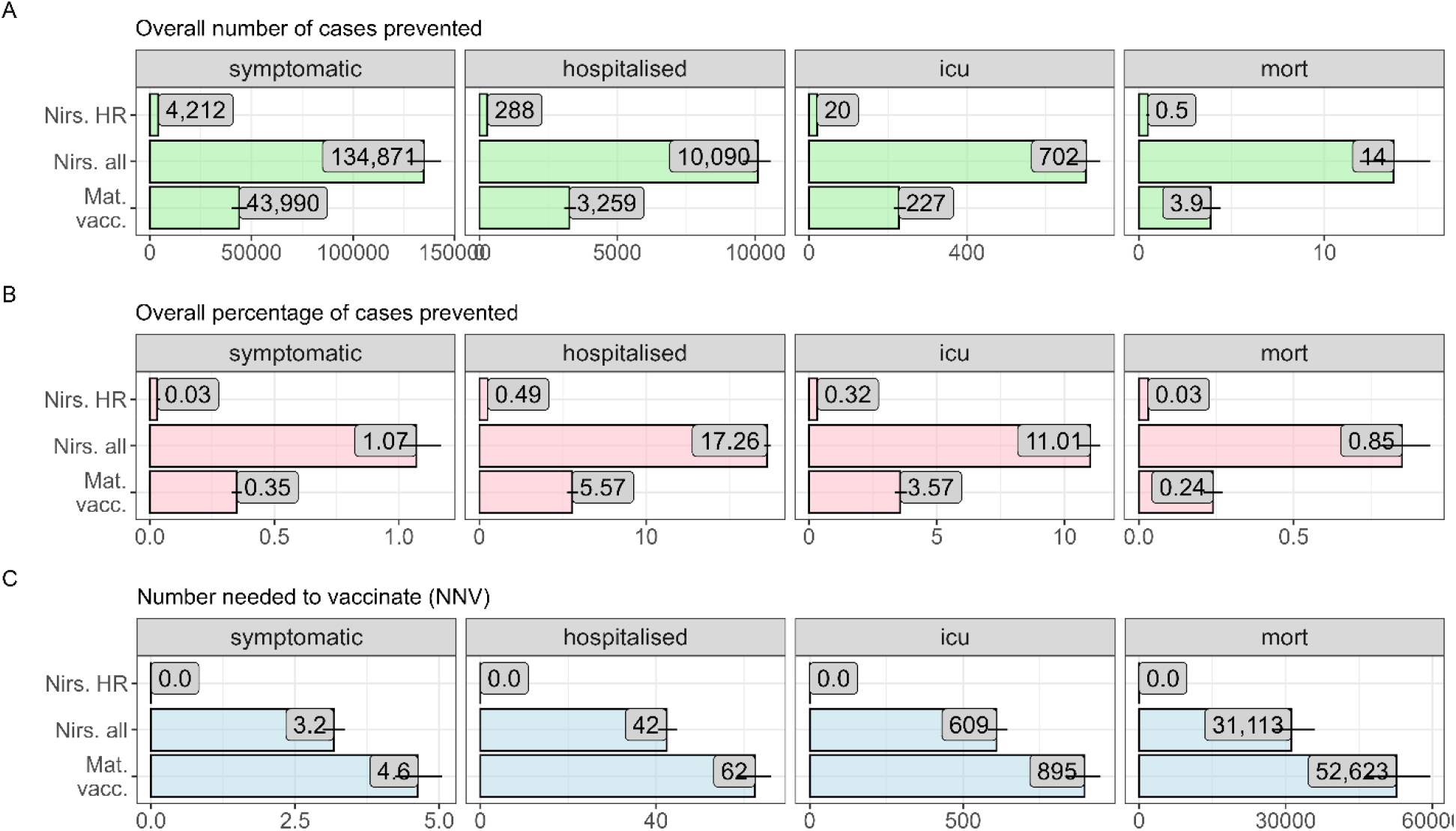
Forward-simulation results for the infant RSV immunisation strategies in Germany over five years compared to the base strategy (Palivizumab for high risk infants). The panels show the overall average number of cases prevented (A), the percentage of cases prevented (B) and the number needed to vaccinate (NNV) (C). The numbers represent the prediction medians, the horizontal lines the 95% prediction interval.

Expanding the eligibility for Nirsevimab to all infants aged 1-5 months with an assumed uptake of 70% (strategy 2) prevents a median of 10,090 (95% PI 9556 - 10561) RSV-specific hospitalisations annually as well as 702 (95% PI 663-738) ICU cases compared to the current standard of care (Fig. 3A). This corresponds overall to a relative reduction of median 17.3% (95% PI 17.1-17.5) of all hospitalisations and ICU admissions and 39.3% (95% PI 39.1-39.5%) of hospitalisations and ICU admissions in infants <1 year. Under this strategy, 42 (95% 41-48) infants would need to be immunised in addition to how many are immunised in the current strategy to prevent one hospitalisation (number needed to vaccinate, NNV), and 609 (95% PI 579-645) to prevent one ICU case (Figure 3C).

Seasonal vaccination of pregnant women with an assumed uptake of 40% (strategy 3) was estimated to prevent on average 3,259 (95% PI 3,075 – 3,489) hospitalisations per year, and 227 (95% PI 215-245) ICU cases (Fig. 3A), with a NNV of 63 (95% PI 58-66) per prevented hospitalisation and 895 (832-948) per ICU case. Overall, this corresponds to a relative reduction of 5.6% of all hospitalisations (95% PI 5.3-5.9), and to a relative reduction of 12.9% (95% PI 12.2-13.7%) of hospitalisations and ICU cases in <1 year olds.

For all infant immunisation strategies, the reduction in disease burden resulted mostly from direct protection among immunised individuals with small indirect effects because of the reduced number of symptomatic infected in the population. Furthermore, the effect of the infant immunisation was stable over the 5 year simulation period for any strategy (fig. S16). The prevented number of cases by age group and strategy over the five years are shown in supplementary figures S12-S15.

Our sensitivity analyses showed that the impact of an immunisation strategy changes proportionally with immunisation uptake because of the limited indirect effects (Fig. S17). We also observed that the seasonal Nirsevimab strategy for all infants aged 1-5 months prevents substantially more cases than the seasonal maternal vaccination for the same uptake. This is due to the circumstance, that infants born before the start of the RSV-season and who are within the target age band during the season are still being immunised with mAbs, while these same infants could not be passively immunised with a maternal vaccine during the season (because they are already born before the season). Hence, seasonal immunisation with mAbs always has the advantage of a larger reach than seasonal maternal vaccination for any uptake level. We also found that year-round maternal vaccination could prevent additional RSV burden but that it has a reduced efficiency (i.e. a higher NNV) compared to a seasonal programme (Supplementary Figure S18).

### Immunisation strategies to protect older adults

Depending on the assumed under-ascertainment of RSV-related hospital admissions, vaccinating older adults aged 75+ years at 40% uptake in addition to immunising all infants with Nirsevimab at 70% uptake (strategy 5) was estimated to prevent between an annual mean of 2,002 and 3,492 RSV hospitalisations and 319-557 ICU cases additionally in the first five years after introducing the vaccination (Figure 5A). This corresponds to 259-452 additional immunisations needed to prevent one RSV-associated additional hospitalisation, and 1,625-2,840 to prevent one additional ICU case (Figure 5B). Vaccinating all individuals aged 65+ once in the first season followed by vaccinating all individuals turning 65 in the following seasons (in addition to Nirsevimab for all infants, strategy 4) prevented slightly more cases (2802-4864) but also requires a larger NNV to prevent one additional hospitalisation (339-589). Overall, we found that the impact and efficiency of the RSV immunisation programme decreases substantially in these age groups (Figure 5A/B) due to a roughly 4 times higher burden of disease in 75+ compared to 55-64-year-old. The absolute reduction in disease burden through vaccination in older adults is largely explained through direct protection in the immunised individuals although there is also a small reduction in disease burden in other non-immunised age-groups when a large share of the older adult population is protected from symptomatic disease during the first simulated years (Figs. S19-S22). The overall reduction of disease burden from the simulated one-time vaccination strategy is mostly driven by effects in the first simulation year when the largest share in the older adult population is protected. It drops substantially in simulation years 3-5 when only individuals who newly age into the target age group are vaccinated (Fig. S16). A repeated seasonal vaccination after a specified time would lead to a larger decrease in the burden, but this was not modelled here since there is no safety or efficacy data on booster vaccination.

**Figure 4:**
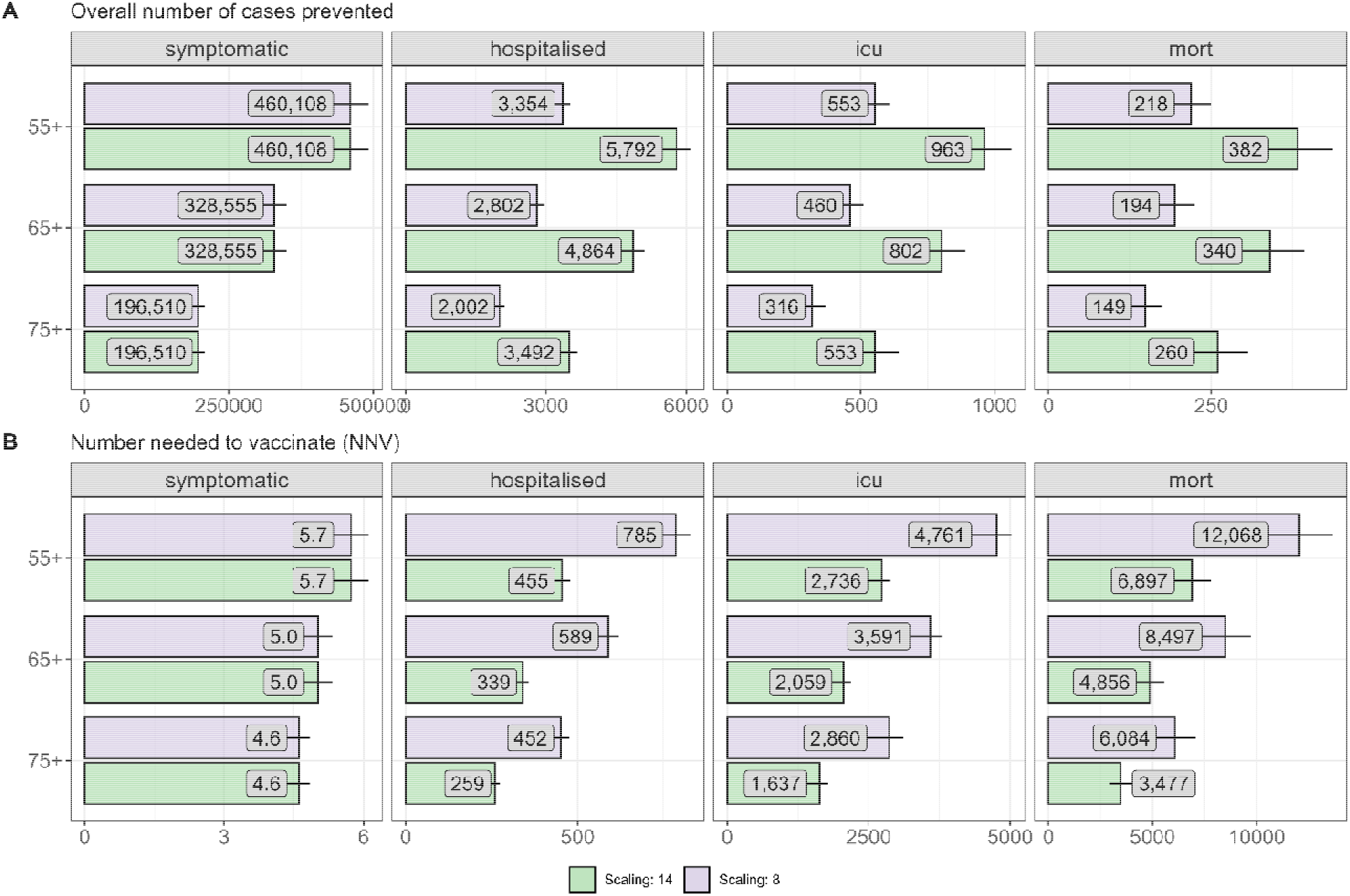
Forward-simulation results for the single dose vaccination of older adults over five years in addition to Nirsevimab for all infants, compared to a base strategy of no adult vaccination (only Nirsevimab for all infants). The panels represent the annual average number of prevented symptomatic cases (A), the percentages prevented (B) and the corresponding numbers needed to vaccinate (C). The numbers represent the prediction medians, the horizontal lines represent the 95% prediction intervals. Under-ascertainment of adult RSV-hospitalisations was accounted for by scaling the RSV-specific hospitalisations with two different scaling factors: 8 and 14 (purple/green).

## Discussion

This study estimated the potential impact of the recently-licensed RSV immunisation products on the RSV disease burden in Germany. We found that changing the current programme for high-risk infants from a short-acting, multi-dose mAB (Palivizumab) to a long-acting, single-dose mAB (Nirsevimab) would reduce the burden of disease in addition to the logistical benefits of administering only a single dose. Expanding eligibility for Nirsevimab to all infants aged 1-5 months prevented a further 18% of RSV-related hospitalisations annually, despite limited indirect effects of immunisation. However, compared to other childhood vaccines, the number needed to immunise to prevent a single death was high due to the relatively low mortality of RSV in this age group. Maternal vaccination would need to achieve a much larger uptake than Nirsevimab to have an impact comparable to Nirsevimab for all infants. We also found that in older adults vaccination in addition to Nirsevimab for all infants could prevent an additional substantial disease burden, although the precise extent of that burden in Germany is not well defined due to under-ascertainment. One of the main drivers of the differences in the projected impact of the strategies for RSV prevention in infants was the assumption that administration of Nirsevimab to all infants would achieve substantially higher coverage than maternal vaccination. Recent experience in Spain has shown an average uptake of 72% ^24^, which suggests our assumption may have been realistic. Higher coverage of maternal vaccination for RSV may be achieved that could substantially reduce the difference in impact compared to mAbs.

The passive protection provided to infants largely disappears within 6-8 months, requiring administration just before the RSV season to optimise the impact. This is particularly challenging for a maternal vaccination programme as the anticipated duration of immunity is shorter than with Nirsevimab. In the UK, seasonal administration was deemed too challenging logistically, for Germany there are no preferences yet. As observed during the COVID-19 pandemic, RSV seasonality can also be disturbed by external factors ^25,26^, which potentially pose additional challenges for a seasonal RSV programme.

While we found that RSV vaccination in older adults could be effective, there is much uncertainty in the underlying disease burden. RSV-associated hospitalisations were estimated based on ICD10 codes with RSV-specific case definitions and then adjusted for under-reporting based on the assumption that in older adults only one in 8 to 14 RSV cases was diagnosed as such during the time period of the empirical data that were used. Moreover, there is residual uncertainty with regards to the duration of protection from the vaccines in this age group. Recent evidence suggests that a substantial amount of the protection is carried over into the second season, ^27^ which could make vaccine administration less time sensitive. However, the number of doses needed has not been determined yet.

The modelling framework allowed us to synthesise diverse pieces of evidence on the epidemiology of RSV in Germany and reflect its uncertainty while quantifying the likely impact of different immunisation strategies. The project was conceived with sufficient lead time to allow for regular stakeholder input into model design and parameterisation. In 2024, the model has been used as part of the evidence-gathering process for the STIKO deliberations on RSV immunisation strategies.

In our analyses, we focused solely on the benefits of vaccination without considering potential risks. While all products assessed here have been licensed and thus deemed by regulatory authorities safe and effective to use, an increase in premature delivery has been reported from Phase III trial after maternal vaccination. The increase was observed only in low- and middle-income countries (i.e. not in high-income countries), and there was no observed increase in mortality. Nevertheless, this may support a more cautious approach e.g. by restricting its use to later in the pregnancy, as recommended in the USA ^28^. Additional studies will be required to assess the validity of the safety signal and to quantify the risk. Our estimates of the benefits can be considered conservative as we did not take into account secondary benefits that may arise from an infant RSV immunisation programme, e.g. through the preservation of scarce bed capacity in paediatric intensive care units ^29^ or the prevention of secondary bacterial pneumonia or antibiotic prescribing ^30^. We also did not consider immunisation of infants in the first 30 days of life due to the current late immunisation with Palivizumab. Including immunisation immediately after birth will likely have an even larger impact on the prevention of cases. The model did not account explicitly for older individuals in high-risk groups or residents in long-term care facilities, who could also be considered a specific target group for vaccination to vaccination against other respiratory pathogens ^31^. Lastly, we assessed the likely impact of RSV immunisation in Germany with the underlying assumption that as of 2024 RSV epidemiology will have largely reverted to pre-pandemic dynamics. As in many other countries, RSV epidemiology in Germany has been heavily perturbed by the Covid19 pandemic. In addition, RSV became notifiable only as of July 2023, making it difficult to interpret post-pandemic changes in RSV incidence due to the enhanced sensitivity of surveillance. However, a number of countries have reported return to pre-pandemic RSV epidemiology in the last year, which supports our modelling assumption of regular RSV dynamics ^32^.

## Conclusions

We find that all three newly available immunisation products against RSV disease have substantial scope to improve health in Germany. The relative merits of the products will depend on yet-uncertain factors, including achievable uptake, the ability to overcome logistical challenges in seasonal administration, the exact burden of disease and the cost of the products.

## Data Availability

This study uses claims data from a German health insurance company that are subject to strict data protection rules according to the German Code of Social Law (“SGB V”). Therefore, these data cannot be made publicly accessible. Public data of the sentinel surveillance can be found online at https://influenza.rki.de. Public data on intensive care stays and in-hospital mortality are available online at https://datenbrowser.inek.org. The programming code for the model and other, public data are available at https://github.com/fkrauer/RSV-VACC-DE.

## Declarations

### Ethics approval and consent to participate

Due to the aggregated nature of the data, no personal patient information was disclosed and no informed consent or ethics approval was necessary for this study.

### Availability of data and materials

This study uses claims data from a German health insurance company that are subject to strict data protection rules according to the German Code of Social Law (SGB V). Therefore, these data cannot be made publicly accessible. Public data of the sentinel surveillance can be found online at https://influenza.rki.de. Public data on intensive care stays and in-hospital mortality are available online at https://datenbrowser.inek.org. The programming code for the model and other, public data are available at https://github.com/fkrauer/RSV-VACC-DE.

### Authors’ contributions

FK: Conceptualization, Methodology, Software, Validation, Formal analysis, Investigation, Writing - Original Draft, Writing - Review & Editing, Visualization, Project administration

FG: Methodology, Software, Validation, Formal analysis, Investigation, Writing - Original Draft, Writing - Review & Editing, Visualization

MTS: Formal analysis, Investigation, Writing - Review & Editing

VS: Methodology, Writing - Review & Editing, Project administration

MK: Methodology, Writing - Review & Editing

MJ: Methodology, Writing - Review & Editing

DH: Methodology, Resources, Writing - Review & Editing US: Resources, Investigation, Writing - Review & Editing

OW: Conceptualization, Methodology, Writing - Review & Editing, Supervision, Funding acquisition

TH: Conceptualization, Methodology, Writing - Review & Editing, Supervision, Project administration, Funding, acquisition

FGS: Methodology, Investigation, Writing - Original Draft, Writing - Review & Editing, Supervision, Project administration

SF: Conceptualization, Methodology, Formal analysis, Investigation, Writing - Original Draft, Writing - Review & Editing, Visualization, Supervision, Project administration

## Acknowledgements

The authors thank the members of the working group on RSV at the German National Immunisation Technical Advisory Group called ‘Standing Committee on Vaccination’ (STIKO) as well as the STIKO Secretariat at RKI for helpful comments. We thank Ulrich Reinacher (RKI) for the technical support in getting the dynamic transmission model running at RKI. We are grateful for all staff and colleagues who tirelessly continue collecting the hospital and sentinel surveillance data. The views expressed in this paper are exclusively those of the authors.

