## supplementary material for "Effectiveness and efficiency of immunisation strategies to prevent RSV among infants and older adults in Germany: a modelling study"

### Methods

#### Model structure

The model is a 4-dimensional object stratified by age, level of infection, epidemiological state and immunisation arm. The arms represent of 6 mutually exclusive infection-immunisation states:

1. non-immunised/unvaccinated individuals
2. infants immunised with mAbs
3. vaccinated individuals
4. Infants immunised through maternal vaccination
5. unvaccinated pregnant women in the third trimester
6. vaccinated pregnant women in the third trimester

We differentiate between active immunisation (vaccination, arm 3) and passive immunisation (mABs, maternal vaccination, arms 2,4). All arms have the same structure of age and infection levels as the no RSV intervention arm (1). The immunisation arms (2,3,4,6) include only the compartments M1, M2, S, E, I, A, and a waning compartment W. The waning compartment allows for Erlang distributed duration of protection. Without immunisation, individuals exists in arm 5 (pregnant women in the third trimester) or arm 1 (the rest). The rate at which individuals move into stratum 5 (third trimester pregnancy) is given by the mean number of daily births, b, the rate of leaving stratum 5 is given by the duration of the third trimester $t_3^{-1}$ (90 days).

We model the immunisation by shifting the proportion to be immunised from arm 1 to the same state, level and age group of the immunisation arm (2 when immunised with mAbs, 3 when vaccinated and not pregnant, 6 when vaccinated an pregnant) (see fig. 1 main text and fig. S0).

We assume that neither passive nor active immunisation prevents infection but reduces the risk of symptomatic disease and hospitalisation, parametrised in line with observed efficacies of the mABs ($PE_s$, $PE_h$) and the vaccine ($VE_s$, $VE_h$) in the Phase 3 trials. Some individuals experience a breakthrough infection in the vaccinated arm and progress to the symptomatic state (*I*) with a probability $p_s * (1 - XE_s)$, where XE is the efficacy for mABs (PE) or the vaccine (VE). The rest progresses to the asymptomatic state. After clearance of infection the immunised individuals return to the immune compartment of the unvaccinated arm with temporary, but full immunity due to the breakthrough infection. Those immunised individuals who were not infected experience a decline of protection and progress to waning compartment W at rate $2 * \omega_X^{-1}$. From the waning compartment, passively immunised proceed at a rate $2 * \omega_X^{-1}$ back to the susceptible state of the unvaccinated arm of the same level because they do not gain immune memory. Actively immunised (=vaccinated) individuals proceed to the susceptible state of the unvaccinated arm of the next level due to gain of immune memory.

Fig. S0. Schematic of flow of individuals in the maternal vaccination strategy.

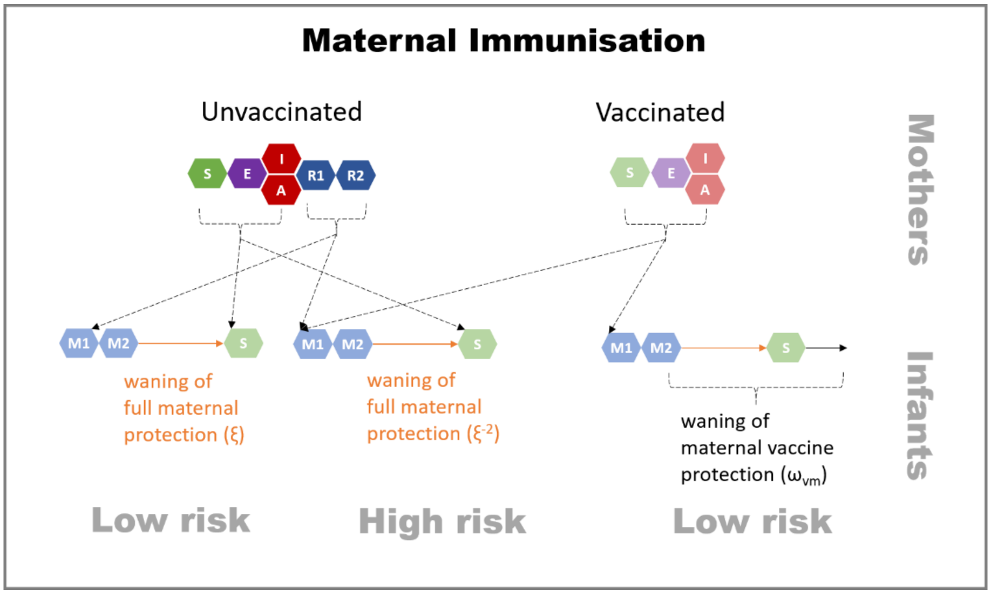

#### Model initialisation and initial conditions

The model was integrated with a non-stiff solver (Tsit5) at tolerances of 1e-4 and the output was returned at daily time steps. The population in each age group was initialised with numbers based on the predictions for 2019 by the Federal Statistical Office for Germany [1]. The initial conditions were chosen such that the whole population in their corresponding age groups were considered susceptible (S) and unvaccinated (arm 1) and the disease was introduced with n=100 infectious individuals in the age group of the 1-year old (see also table S1). All the other states were considered zero. The model was then simulated for 20 years until the disease dynamics reached a stable seasonality (stability was checked visually after fitting). The states after 20 years of simulation were then used as starting points for both the fitting process and the vaccination simulation, the 20-year run-in phase was discarded.

Table S1. Initial states by compartment, level and age group

| Age group | Compartment | N high risk (level 0) | N low risk (level 1) |
|---|---|---|---|
| 0 months | S | 2247.12 | 62325.377 |
| 1 month | S | 2247.12 | 62325.377 |
| 2 months | S | 2247.12 | 62325.377 |
| 3 months | S | 2247.12 | 62325.377 |
| 4 months | S | 2247.12 | 62325.377 |
| 5 months | S | 2247.12 | 62325.377 |
| 6 months | S | 2247.12 | 62325.377 |
| 7 months | S | 2247.12 | 62325.377 |
| 8 months | S | 2247.12 | 62325.377 |
| 9 months | S | 2247.12 | 62325.377 |
| 10 months | S | 2247.12 | 62325.377 |
| 11 months | S | 2247.12 | 62325.377 |
| 1 year | S | 0.0 | 794032.0 |
| 1 year | E | 0.0 | 100.0 |
| 2 years | S | 0.0 | 802,415 |
| 3 years | S | 0.0 | 807,816 |
| 4 years | S | 0.0 | 782,143 |
| 5-9 years | S | 0.0 | 3,730,000 |
| 10-14 years | S | 0.0 | 3,700,000 |
| 15-24 years | S | 0.0 | 8,550,000 |
| 25-34 years | S | 0.0 | 10,600,000 |
| 35-44 years | S | 0.0 | 10,200,000 |
| 45-54 years | S | 0.0 | 12,000,000 |
| 55-64 years | S | 0.0 | 12,400,000 |
| 65-74 years | S | 0.0 | 8,530,000 |
| 75+ years | S | 0.0 | 9,560,000 |
| **Subtotal** | | **26965.476** | **83,139,745.524** |
| **Total** | | | **83,166,711** |

### Model equations

#### Transmission model

Table S2. Equations of the base model structure including transmissions and epidemiological transitions. Shaded rows indicate states that do not exist at the given level of re-infection.

| State | level | Births and ageing in | ageing out | deaths | Transmissions | Transitions |
|---|---|---|---|---|---|---|
| $M1_0^a$ | 0 | $b * p_h * p_m$ | $-\varepsilon^a * M1_0^a$ | $- \mu_{0,M1}^a$ | | $-2 * (z * \xi)^{-1} * M1_0^a$ |
| $M2_0^a$ | 0 | $\varepsilon^{a-1} * M2_0^{a-1}$ | $-\varepsilon^a * M2_0^a$ | $- \mu_{0,M2}^a$ | | $2 * (z * \xi)^{-1} * M1_0^a - 2 * (z * \xi)^{-1} * M2_0^a$ |
| $S_0^a$ | 0 | $b * p_h * (1 - p_m)$ | $-\varepsilon^a * S_0^a$ | $-\mu_{0,S}^a$ | $-\lambda^a * \delta_0 * S_0^a$ | $2 * (z * \xi)^{-1} * M2_0^a$ |
| $E_0^a$ | 0 | $\varepsilon^{a-1} * E_0^{a-1}$ | $-\varepsilon^a * E_0^a$ | $-\mu_{0,E}^a$ | $\lambda^a * \delta_0 * S_0^a$ | $-\sigma * E_0^a$ |
| $I_0^a$ | 0 | $\varepsilon^{a-1} * I_0^{a-1}$ | $-\varepsilon^a * I_0^a$ | $-\mu_{0,I}^a$ | | $\sigma * p_s * E_0^a - \gamma_0 * I_0^a$ |
| $A_0^a$ | 0 | $\varepsilon^{a-1} * A_0^{a-1}$ | $-\varepsilon^a * A_0^a$ | $-\mu_{0,A}^a$ | | $\sigma * (1 - p_s) * E_0^a - \gamma_0 * A_0^a$ |
| $R1_0^a$ | 0 | $\varepsilon^{a-1} * R1_0^{a-1}$ | $-\varepsilon^a * R1_0^a$ | $-\mu_{0,R1}^a$ | | $\gamma_0 * I_0^a + \gamma_0 * A_0^a - 2 * \omega^{-2} * R1_0^a$ |
| $R2_0^a$ | 0 | $\varepsilon^{a-1} * R2_0^{a-1}$ | $-\varepsilon^a * R2_0^a$ | $-\mu_{0,R2}^a$ | | $2 * \omega^{-2} * R1_0^a - 2 * \omega^{-2} * R2_0^a$ |
| $M1_1^a$ | 1 | $b * (1 - p_h) * p_m$ | $-\varepsilon^a * M1_1^a$ | $-\mu_{1,M1}^a$ | | $-2 * \xi^{-1} * M1_1^a$ |
| $M2_1^a$ | 1 | $\varepsilon^{a-1} * M2_1^{a-1}$ | | $-\mu_{0,M2}^a$ | | $2 * \xi^{-1} * M1_1^a - 2 * \xi^{-1} * M2_1^a$ |
| $S_1^a$ | 1 | $b * (1 - p_h) * (1 - p_m)$ | $-\varepsilon^a * S_1^a$ | $-\mu_{1,S}^a$ | $-\lambda^a * \delta_1 * S_1^a$ | $2 * \xi^{-1} * M2_1^a + 2 * \omega^{-2} * R2_0^a$ |
| $E_1^a$ | 1 | $\varepsilon^{a-1} * E_1^{a-1}$ | $-\varepsilon^a * E_1^a$ | $-\mu_{1,E}^a$ | $\lambda^a * \delta_1 * S_1^a$ | $-\sigma * E_1^a$ |
| $I_1^a$ | 1 | $\varepsilon^{a-1} * I_1^{a-1}$ | $-\varepsilon^a * I_1^a$ | $-\mu_{1,I}^a$ | | $\sigma * p_s * E_1^a - \gamma_1 * I_1^a$ |
| $A_1^a$ | 1 | $\varepsilon^{a-1} * A_1^{a-1}$ | $-\varepsilon^a * A_1^a$ | $-\mu_{1,A}^a$ | | $\sigma * (1 - p_s) * E_1^a - \gamma_1 * A_1^a$ |
| $R1_1^a$ | 1 | $\varepsilon^{a-1} * R1_1^{a-1}$ | $-\varepsilon^a * R1_1^a$ | $-\mu_{1,R1}^a$ | | $\gamma_1 * I_1^a + \gamma_1 * A_1^a - 2 * \omega^{-2} * R1_1^a$ |
| $R2_1^a$ | 1 | $\varepsilon^{a-1} * R2_1^{a-1}$ | $-\varepsilon^a * R2_1^a$ | $-\mu_{1,R2}^a$ | | $2 * \omega^{-2} * R1_1^a - 2 * \omega^{-2} * R2_1^a$ |
| $M1_2^a$ | 2 | | | | | |
| $M2_2^a$ | 2 | | | | | |
| $S_2^a$ | 2 | $\varepsilon^{a-1} * S_2^{a-1}$ | $-\varepsilon^a * S_2^a$ | $-\mu_{2,S}^a$ | $-\lambda^a * \delta_2 * S_2^a$ | $2 * \omega^{-2} * R2_1^a$ |
| $E_2^a$ | 2 | $\varepsilon^{a-1} * E_2^{a-1}$ | $-\varepsilon^a * E_2^a$ | $-\mu_{2,E}^a$ | $\lambda^a * \delta_2 * S_2^a$ | $-\sigma * E_2^a$ |
| $I_2^a$ | 2 | $\varepsilon^{a-1} * I_2^{a-1}$ | $-\varepsilon^a * I_2^a$ | $-\mu_{2,I}^a$ | | $\sigma * p_s * E_2^a - \gamma_2 * I_2^a$ |
| $A_2^a$ | 2 | $\varepsilon^{a-1} * A_2^{a-1}$ | $-\varepsilon^a * A_2^a$ | $-\mu_{2,A}^a$ | | $\sigma * (1 - p_s) * E_2^a - \gamma_2 * A_2^a$ |
| $R1_2^a$ | 2 | $\varepsilon^{a-1} * R1_2^{a-1}$ | $-\varepsilon^a * R1_2^a$ | $-\mu_{2,R1}^a$ | | $\gamma_2 * I_2^a + \gamma_2 * A_2^a - 2 * \omega^{-2} * R1_2^a$ |
| $R2_2^a$ | 2 | $\varepsilon^{a-1} * R2_2^{a-1}$ | $-\varepsilon^a * R2_2^a$ | $-\mu_{2,R2}^a$ | | $2 * \omega^{-2} * R1_2^a - 2 * \omega^{-2} * R2_2^a$ |
| $M1_3^a$ | 3 | | | | | |
| $M2_3^a$ | 3 | | | | | |

| | | | | | | |
|---|---|---|---|---|---|---|
| $S_3^a$ | 3 | $\varepsilon^{a-1} * S_3^{a-1}$ | $-\varepsilon^a * S_3^a$ | $-\mu_{3,S}^a$ | $-\lambda^a * \delta_3 * S_3^a$ | $2 * \omega^{-2} * R2_2^a$ |
| $E_3^a$ | 3 | $\varepsilon^{a-1} * E_3^{a-1}$ | $-\varepsilon^a * E_3^a$ | $-\mu_{3,E}^a$ | $\lambda^a * \delta_3 * S_3^a$ | $-\sigma * E_3^a$ |
| $I_3^a$ | 3 | $\varepsilon^{a-1} * I_3^{a-1}$ | $-\varepsilon^a * I_3^a$ | $-\mu_{3,I}^a$ | | $\sigma * p_s * E_3^a - \gamma_3 * I_3^a$ |
| $A_3^a$ | 3 | $\varepsilon^{a-1} * A_3^{a-1}$ | $-\varepsilon^a * A_3^a$ | $-\mu_{3,A}^a$ | | $\sigma * (1 - p_s) * E_3^a - \gamma_3 * A_3^a$ |
| $R1_3^a$ | 3 | $\varepsilon^{a-1} * R1_3^{a-1}$ | $-\varepsilon^a * R1_3^a$ | $-\mu_{3,R1}^a$ | | $\gamma_3 * I_3^a + \gamma_3 * A_3^a - 2 * \omega^{-2} * R1_3^a$ |
| $R2_3^a$ | 3 | $\varepsilon^{a-1} * R2_3^{a-1}$ | $-\varepsilon^a * R2_3^a$ | $-\mu_{3,R2}^a$ | | $2 * \omega^{-2} * R1_3^a - 2 * \omega^{-2} * R2_3^a$ |
| $M1_4^a$ | 4 | | | | | |
| $M2_4^a$ | 4 | | | | | |
| $S_4^a$ | 4 | $\varepsilon^{a-1} * S_4^{a-1}$ | $-\varepsilon^a * S_4^a$ | $-\mu_{4,S}^a$ | $-\lambda^a * \delta_4 * S_4^a$ | $2 * \omega^{-2} * R2_3^a + 2 * \omega^{-2} * R2_4^a$ |
| $E_4^a$ | 4 | $\varepsilon^{a-1} * E_4^{a-1}$ | $-\varepsilon^a * E_4^a$ | $-\mu_{4,E}^a$ | $\lambda^a * \delta_4 * S_4^a$ | $-\sigma * E_4^a$ |
| $I_4^a$ | 4 | $\varepsilon^{a-1} * I_4^{a-1}$ | $-\varepsilon^a * I_4^a$ | $-\mu_{4,I}^a$ | | $\sigma * p_s * E_4^a - \gamma_3 * I_3^a$ |
| $A_4^a$ | 4 | $\varepsilon^{a-1} * A_4^{a-1}$ | $-\varepsilon^a * A_4^a$ | $-\mu_{4,A}^a$ | | $\sigma * (1 - p_s) * E_4^a - \gamma_4 * A_4^a$ |
| $R1_4^a$ | 4 | $\varepsilon^{a-1} * R1_4^{a-1}$ | $-\varepsilon^a * R1_4^a$ | $-\mu_{4,R1}^a$ | | $\gamma_4 * I_4^a + \gamma_4 * A_4^a - 2 * \omega^{-2} * R1_4^a$ |
| $R2_4^a$ | 4 | $\varepsilon^{a-1} * R2_4^{a-1}$ | $-\varepsilon^a * R2_4^a$ | $-\mu_{4,R2}^a$ | | $2 * \omega^{-2} * R1_4^a - 2 * \omega^{-2} * R2_4^a$ |

#### Immunisation model

The immunisation strategy simulation model has an additional waning compartment (W) in each age group and each level of reinfection (except for the Palivizumab immunisation in the fitted model). The tables below only show the terms that differ from the base transmission model or that are additional to the base transmission model. In addition to the term listed below, the arms also have all the demographic terms from the base model (births, ageing, deaths), which were omitted from the table for simplicity. Red terms are instantaneous shifts of individuals between the compartments at the beginning of the intervention time window (=those who are in the eligible age groups at the start of the intervention). Blue terms are continuous shifts of individuals during the intervention time window (= those who newly age into the age groups eligible for intervention). Visibly sick individuals (i.e.in the I state) are not immunised. Green terms denote the waning of protection from the immunization. Purple terms indicate breakthrough infections. A square bracket after the state name indicates, which arm/stratum the state belongs to ([1] = unvaccinated arm, [2] = Palivizumab arm, [3] = vaccinated arm, [4] passively immunized infants through maternal vaccination, [5] = unvaccinated women in the 3rd trimester, [6] = vaccinated women in the 3rd trimester). Shaded cells indicate states that do not exist for the given arm.

#### mAb immunisation (fitted model)

Table S3. Equations of the fitted Palivizumab immunisation model including exponential waning. Infants can technically also be immunised when in the E, I, A or R1/R2 state, but these are considered wasted doses as we assume they do not have any effect, hence they are not modelled.

| State | level | Unvaccinated arm (=1) | mAB vaccinated arm (=2) |
|---|---|---|---|
| $M1_0^a$ | 0 | $-p_p * M1[1]_0^a$ | $p_p * M1[1]_0^a + p_p * \epsilon^{a-1} * M1_0^{a-1}$ |
| $M2_0^a$ | 0 | $-p_p * M2[1]_0^a$ | $p_p * M1[1]_0^a + p_p * \epsilon^{a-1} * M1_0^{a-1}$ |
| $S_0^a$ | 0 | $-p_p * S[1]_0^a + \omega_p^{-1} * S[2]_0^a$ | $p_p * S[1]_0^a + p_p * \epsilon^{a-1} * M1_0^{a-1} - \omega_p^{-1} * S[2]_0^a - \lambda^a * \delta_0 * (1 - PE_I) * S[2]_0^a$ |
| $E_0^a$ | 0 | | $\lambda^a * \delta_0 * (1 - PE_I) * S[2]_0^a - \sigma * E[2]_4^a$ |
| $I_0^a$ | 0 | | $\sigma * p_s * \frac{(1 - PE_S)}{(1 - PE_I)} * E[2]_4^a$ |
| $A_0^a$ | 0 | | $\sigma * \left(1 - \left(p_s * \frac{(1 - PE_S)}{(1 - PE_I)}\right)\right) * E[2]_4^a$ |
| $R1_0^a$ | 0 | $\gamma_0 * I[2]_0^a + \gamma_0 * A[2]_0^a$ | |
| $R2_0^a$ | 0 | | |

#### mAb immunisation (vaccination model)

Table S4. Equations of the Nirsevimab or Palivizumab immunisation in the vaccination model including the Erlang2 waning. The table below only shows the terms for passive mAB immunization of high risk individuals (level 0). If all infants are included in the immunization strategy (strategy 2), the equation terms are identical for the other levels (except for the level subscript). We have omitted these equations for clarity. Infants can technically also be immunised when in the E, I, A or R1/R2 state, but these are considered wasted doses as we assume they do not have any effect, hence they are not modelled.

| State | level | Unvaccinated arm (=1) | mAB vaccinated arm (=2) |
|---|---|---|---|
| $M1_0^a$ | 0 | $-p_p * M1[1]_0^a$ | $p_p * M1[1]_0^a + p_p * \epsilon^{a-1} * M1_0^{a-1}$ |
| $M2_0^a$ | 0 | $-p_p * M2[1]_0^a$ | $p_p * M1[1]_0^a + p_p * \epsilon^{a-1} * M1_0^{a-1}$ |
| $S_0^a$ | 0 | $-p_p * S[1]_0^a + 2 * \omega_p^{-1} * W[2]_0^a$ | $p_p * S[1]_0^a + p_p * \epsilon^{a-1} * M1_0^{a-1} - 2 * \omega_p^{-1} * S[2]_0^a - \lambda^a * \delta_0 * (1 - PE_i) * S[2]_0^a$ |
| $E_0^a$ | 0 | | $\lambda^a * \delta_0 * (1 - PE_I) * S[2]_0^a - \sigma * E[2]_4^a$ |
| $I_0^a$ | 0 | | $\sigma * p_s * \frac{(1 - PE_S)}{(1 - PE_I)} * E[2]_4^a$ |
| $A_0^a$ | 0 | | $\sigma * \left(1 - \left(p_s * \frac{(1 - PE_S)}{(1 - PE_I)}\right)\right) * E[2]_4^a$ |
| $R1_0^a$ | 0 | $\gamma_0 * I[2]_0^a + \gamma_0 * A[2]_0^a$ | |
| $R2_0^a$ | 0 | | |
| $W_0^a$ | 0 | | $2 * \omega_p^{-1} * S[2]_0^a - 2 * \omega_p^{-1} * W[2]_0^a$ |

##### Maternal immunisation (infants)

Table S5. Equations of the maternal immunisation model in infants. Maternally immunised infants can only be born into levels 0 or1 to maternally immunised mothers (arm 4) or non-immunised mothers (arm 1) (burgundy). The parameters $p_{mv}$and $p_m$ are the proportion of infants born to maternally immunised mothers or to mothers immunised by infection, respectively. Both depends on the current number of pregnant women in the arm 5 or 6, which varies seasonally and with vaccination timing. Therefore, these parameters are calculated instantly inside the model, and may vary throughout the season.

| State | level | Unvaccinated arm (=1) | Maternally immunised arm (=4) |
|---|---|---|---|
| $M1_0^a$ | 0 | $b * p_h * (p_m)$ | |
| $S_0^a$ | 0 | $b * p_h * (1 - p_m - p_{mv})$ | |
| $M1_1^a$ | 1 | $b * (1 - p_h) * p_m$ | $b * (1 - p_h) * p_{mv}$ |
| $S_1^a$ | 1 | $b * (1 - p_h) * (1 - p_m - p_{mv}) + 2 * \omega_{vm}^{-1} * W[4]_1^a$ | $-2 * \omega_{vm}^{-1} * S[4]_1^a - \lambda^a * \delta_1 * (1 - VE_{im}) * S[4]_1^a$ |
| $E_1^a$ | 1 | | $\lambda^a * \delta_1 * (1 - VE_{im}) * S[4]_1^a - \sigma * E[4]_1^a$ |
| $I_1^a$ | 1 | | $\sigma * p_s * \frac{(1 - VE_{sm})}{(1 - VE_{im})} * E[4]_1^a$ |
| $A_1^a$ | 1 | | $\sigma * \left(1 - \left(p_s * \frac{(1 - VE_{sm})}{(1 - VE_{im})}\right)\right) * E[4]_1^a$ |
| $R1_1^a$ | 1 | $\gamma_1 * I[4]_1^a + \gamma_1 * A[4]_1^a$ | |
| $R2_1^a$ | 1 | | |
| $W_1^a$ | 1 | | $2 * \omega_{vm}^{-1} * S[4]_1^a - 2 * \omega_{vm}^{-1} * W[4]_1^a$ |

#### Vaccination of pregnant women

Table S6. Equations of the maternal immunisation model in mothers. N[1] denotes the population size in the age groups 25-45 years. The term $b * X[1][_j^a/N[1]$ is number of women who enter the third trimester in pregnancy every day for any epidemiological state X, and any age group a and level j in arm 1 (turquoise). The term $t_3{}^{-1} * X[5/6]_j^a$ is the number of women who give birth every day for any epidemiological state X, and any age group a and level j, in arms 5/6 (orange). At this point, they move either to unvaccinated, not pregnant arm (5 → 1) or to the vaccinated, not-pregnant arm (6 → 3) (equations not shown).

| State | level | Unvaccinated arm (=1) | Pregnant women, unvaccinated (=5) | Pregnant women, vaccinated (=6) |
|---|---|---|---|---|
| $S_j^a$ | $j$ | $t_3{}^{-1} * S[5]_j^a - b * \frac{S[1]_j^a}{N[1]}$ | $b * \frac{S[1]_j^a}{N[1]} - t_3{}^{-1} * S[5]_j^a - v * S[5]_j^a - v * b * \frac{S[1]_j^a}{N[1]}$ | $v * S[5]_j^a + v * b * \frac{S[1]_j^a}{N[1]} - 2 * \omega_v^{-1} * S[6]_j^a$ $-\lambda^a * \delta_1 * (1 - VE_i) * S[6]_j^a - t_3{}^{-1} * S[6]_j^a$ |
| $E_j^a$ | $j$ | $t_3{}^{-1} * W[5]_j^a - b * \frac{E[1]_j^a}{N[1]}$ | $b * \frac{S[1]_j^a}{N[1]} - t_3{}^{-1} * E[5]_j^a$ | $\lambda^a * \delta_1 * (1 - VE_i) * S[6]_j^a$ |
| $I_j^a$ | $j$ | $t_3{}^{-1} * I[5]_j^a - b * \frac{I[1]_j^a}{N[1]}$ | $b * \frac{S[1]_j^a}{N[1]} - t_3{}^{-1} * I[5]_j^a$ | $\sigma * p_s * \frac{(1 - VE_s)}{(1 - VE_i)} * E[6]_j^a$ |
| $A_j^a$ | $j$ | $t_3{}^{-1} * A[5]_j^a - b * \frac{A[1]_j^a}{N[1]}$ | $b * \frac{A[1]_j^a}{N[1]} - t_3{}^{-1} * A[5]_j^a$ | $\sigma * \left(1 - \left(p_s * \frac{(1 - VE_s)}{(1 - VE_i)}\right)\right) * E[6]_j^a$ |
| $R1_j^a$ | $j$ | $t_3{}^{-1} * R1[5]_j^a - b * \frac{R1[1]_j^a}{N[1]}$ | $b * \frac{R1[1]_j^a}{N[1]} - t_3{}^{-1} * R1[5]_j^a$ | |
| $R2_j^a$ | $j$ | $t_3{}^{-1} * R2[5]_j^a - b * \frac{R2[1]_j^a}{N[1]}$ | $b * \frac{R2[1]_j^a}{N[1]} - t_3{}^{-1} * R2[5]_j^a$ | |
| $W_j^a$ | $j$ | | | $2 * \omega_v^{-1} * S[5]_j^a - 2 * \omega_v^{-1} * W[6]_j^a$ |
| $S_{j+1}^a$ | $j$+1* | | $2 * \omega_v^{-1} * W[6]_j^a$ | |

*if j=4, then j+1 =4, since there are only 4 levels of infection

#### Vaccination of older adults

Table S7. Equations of the older adult immunisation model. Individuals can technically also be vaccinated when in the states E,I,A,R1 or R2, but we assume that this would not lead to additional immunity (only to wasted doses) and hence do not model it in the transmission model.

| State | level | Unvaccinated arm (=1) | Vaccinated arm (=3) |
|---|---|---|---|
| $S_j^a$ | $j$ | $-v * S[1]_j^a$ | $v * S[1]_j^a + v * \epsilon^{a-1} * S[1]_j^{a-1} - 2 * \omega_v^{-1} * S[3]_j^a$ |
| $W_j^a$ | $j$ | | $2 * \omega_v^{-1} * S[3]_j^a - 2 * \omega_v^{-1} * W[3]_j^a$ |
| $S_{j+1}^a$ | j+1* | $2 * \omega_v^{-1} * W[3]_j^a$ | |

*if j=4, then j+1 =4, since there are only 4 levels of infection

#### Outcome variables

Table S8. Equations of the modelled outcomes in all arms (CS = Symptomatic cases, CH = Hospitalised cases, haj = probability of hospitalisation when symptomatic in age group a and level j). Shaded cells indicate states that do not exist for the given arm.

| Arm | Symptomatic cases $CS_j^a$ | Hospitalisations $CH_j^a$ |
|---|---|---|
| Unvaccinated [1] | $\sigma * p_s * E[1]_j^a$ | $CS[1]_j^a * h_j^a$ |
| mAB [2] | $\sigma * p_s * \frac{(1-PE_s)}{(1-PE_i)} * E[2]_j^a$ | $CS[2]_j^a * h_j^a * \frac{(1-PE_h)}{(1-PE_s)}$ |
| Vaccinated [3] | $\sigma * p_s * \frac{(1-VE_s)}{(1-VE_i)} * E[3]_j^a$ | $CS[3]_j^a * h_j^a * \frac{(1-VE_h)}{(1-VE_s)}$ |
| Maternally vaccinated infants [4] | $\sigma * p_s * \frac{(1-VE_{sm})}{(1-VE_{im})} * E[4]_j^a$ | $CS[4]_j^a * h_j^a * \frac{(1-VE_{hm})}{(1-VE_{sm})}$ |
| Pregnant women, unvaccinated [5] | $\sigma * p_s * E[5]_j^a$ | $CS[5]_j^a * h_j^a$ |
| Pregnant women, vaccinated [6] | $\sigma * p_s * \frac{(1-VE_s)}{(1-VE_i)} * E[5]_j^a$ | $CS[6]_j^a * h_j^a * \frac{(1-VE_h)}{(1-VE_s)}$ |

#### Model fitting

The model tracks the cumulative incidence of symptomatic infections (CS) to calculate a) the weekly incidence of reported symptomatic cases in age groups matched to those in the surveillance (AGI) data, and b) the quarterly incidence of reported hospitalised cases in each of the 25 age groups. For (a) we assumed that an age-dependent proportion of cases $q_i$ was detected. Likewise, for (b), we assumed that the probability of hospitalisation $h$ depends on age (*i*) and risk group.

These two model outputs were used to fit the model to the AGI surveillance and the TK hospitalisation data. Each output was fitted twofold: a) The total cases at each time point (summed over all age groups) and b) The proportions of the cases in the individual age groups (summed over time). The first was fitted with a Negative-Binomial likelihood with a dispersion parameter $\psi$, the second was fitted with a Multinomial likelihood. Additionally, we calculated the proportion of individuals <1 year at levels 0 and 1, who were not yet in the R states (sero-negative) and fit them to the proportion of sero-negatives from the Pienter study with monthly age groups assuming a binomial likelihood. The total log likelihood was calculated as the sum of the individual log likelihoods.

The model was fitted using a No-U-Turn Sampler (NUTS) implemented in the Julia package AdvancedHMC [2] with 4 chains with a burn-in of 500 samples and 1000 accepted samples per chain.

### Parameters

Table S9. Demographic parameters

| Symbol | Parameter | Fixed value / Prior | Ref |
|---|---|---|---|
| $\epsilon^a$ | Transition rate out of age group a (=1/duration of age group a in days) | varying | See below |
| $\mu_{j,k}^a$ | age (a)-, level (j)- and state (k) specific mortality rates | varying | See below |
| b | Daily births | 2132 | [1] |
| $i_m$ | Index of child-bearing age groups | 21:22 (25-45 years) | |
| $p_h$ | Proportion of children born as high risk | 0.0348 | See below |
| $p_m$ | Proportion of infants born with maternal immunity (= into M1) | Varying, depends on proportion of pregnant in the R compartments | See below |

| | | |
|---|---|---|
| $t_3$ | Duration of third trimester | 90 days |

Table S10. Parameters of transmission and disease (bold parameters are estimated)

| Symbol | Parameter | Fixed value / Prior | Ref |
|---|---|---|---|
| $p_s$ | Age-dependent proportion of symptomatic cases among all cases | Varying (0.0, 1.0)<br>$p_s$ in high risk infants 0-12 months is assumed to be 1.0. | see below |
| ϕ | Phase shift of seasonal transmission rate | 0.5467033 | See below |
| σ | Duration of latent period (days) $^{-1}$ | 4.0 | [3] |
| α | Reduction of infectiousness of asymptomatics | 0.2 | [4] |
| $\gamma_0$ | Duration of infectiousness during first infection | 9.0 | [5] |
| g | Proportional reduction of duration of infectiousness at each level of re-infection | 0.74 | see below |
| z | Reduction of duration of maternal immunity in high risk infants | 0.5 | [6] |
| δ0 | Proportional reduction of susceptibility at first infection (high risk) | 1.0 | |
| δ1 | Proportional reduction of susceptibility at first infection (low risk) | 1.0 | |
| δ2 | Proportional reduction of susceptibility at second infection | 1.0 | [7] |
| **δ3** | Proportional reduction of susceptibility at third infection | Uniform(0.0, 1.0) | |

| | | |
|---|---|---|
| **δ4** | Proportional reduction of susceptibility at fourth and subsequent infection | Uniform(0.0, 1.0) |
| **β** | Average transmission rate | Beta(2.5, 5.0) |
| **η** | Amplitude of transmission rate | Beta(1.0, 1.0) |
| [illegible] | Average duration of immunity (days) | truncated(Gamma(14.0, 16.0), 100.0, 365.0) |
| **ξ** | Average duration of maternal immunity (days) | truncated(Gamma(2.5, 30.0), 10.0, 180.0) |

Table S11. Parameters related to the reporting of symptomatic cases and hospitalised cases (bold parameters are estimated)

| | | | |
|---|---|---|---|
| $k_h$ | Increased risk of hospitalization of high risk vs low risk babies | 3.0 | [8] |
| q | Age-dependent detection rate of symptomatic cases | varying | see below |
| h | Age-dependent reported hospitalisation probability in symptomatic 1+ year olds | varying | See below |
| [illegible]$_1$ | Scaling of q | truncated(Normal(1.0, 0.2), 0.5, 1.5) | See below |
| [illegible]$_2$ | Scaling of h | truncated(Normal(1.0, 0.2), 0.5, 1.5) | See below |
| **$a_1$** | Lower bound of exponential decay model for hospitalisation risk in <1 year old high risk individuals | Uniform(0.01, 0.2) | See below |
| **$a_2$** | Upper bound of exponential decay model for hospitalisation risk in <1 year old high risk individuals | Uniform(0.2, 3.0) | See below |

| | | | |
|---|---|---|---|
| $a_3$ | rate of exponential decay model for hospitalisation risk in <1 year old high risk individuals | Uniform(0.0, 5.0) | See below |
| ψ | Dispersion parameter of NegBin | 0.02 | |

Table S12. Parameters of immunisation

| Symbol | Parameter | Clinical trial estimate | Model estimate |
|---|---|---|---|
| | **Palivizumab** | | |
| $PE_i$ | Effectiveness against infection | | 0.0 |
| $PE_s$ | Effectiveness against ARI | 0.67 [9] during 30 days | 0.82 |
| $PE_h$ | Effectiveness against hospitalisation | 0.78 [10] during 30 days | 0.95 |
| $\omega_p$ | Duration of protection (days) | | 40 days |
| $p_p$ | Immunisation uptake (proportion) | | 0.9 |
| | **Nirsevimab** | | |
| $PE_i$ | Effectiveness against infection | | 0.0 |
| $PE_s$ | Effectiveness against ARI | 0.67 (assumption) | 0.96 |
| $PE_h$ | Effectiveness against hospitalisation | 0.784 [11] during 150 days | 1.00 |
| $\omega_p$ | Duration of protection (days) | | 160 |
| $p_p$ | Immunisation uptake (proportion) | | 0.7 or 0.9 |
| | **Maternal vaccine** | | |

| | | | |
|---|---|---|---|
| $VE_i$ | Effectiveness against infection | | Mothers: 0.0<br>Infants: 0.0 |
| $VE_s$ | Effectiveness against ARI | Mothers: 0.621 [12] during 212 days<br>Infants: 0.571 [13] during 180 days | Mothers: 0.67<br>Infants: 0.75 |
| $VE_h$ | Effectiveness against hospitalisation | Mothers: 0.857 [12]<br>Infants: 0.694 [13] | Mothers: 0.92<br>Infants: 0.75 |
| $\omega_v$, $\omega_{vm}$ | Duration of protection (days) | | Mothers: 712 days<br>Infants: 200 days |
| v | Immunisation uptake (proportion) | | 0.4 |
| $p_{mv}$ | Proportion of infants born with maternal protection due to maternal vaccination (= into M1) | Varying, depends on the proportion of pregnant women who are vaccinated | |
| | **Older adult vaccine** | | |
| $VE_i$ | Effectiveness against infection | | 0.0 |
| $VE_s$ | Effectiveness against ARI | 0.717 [14] during 203 days | 0.77 |
| $VE_h$ | Effectiveness against hospitalisation | 0.941 [14] during 203 days | 1.00 |
| $\omega_v$ | Duration of protection (days) | | 712 days |
| v | Immunisation coverage (proportion) | | 0.4 |

#### Proportion of infants in the high risk group ($p_h$)

The high risk group is defined as infants that are born pre-term (<36 gestational week), that have bronchopulmonary dysplasia (BHP, which are mostly pre-term born infants) or congenital heart disease (CHD). The proportion of infants born into the high risk group $p_h$ was estimated based on two proportions. The proportion of infants born pre-term was taken from literature as 0.026 [15]. The overall proportion of infants born with CHD in literature is 0.01076, and the probability of CHD among pre-term is 0.187 [16]. The probability *x* of CHD among term born infants was then estimated as

$$0.026 * 0.187 + (1 - 0.026) * x = 0.01076$$

$$x = \frac{0.01076 - 0.026 * 0.187}{(1 - 0.026)} = 0.0088$$

The total proportion of infants born in the high risk group is the sum of the proportion pre-terms and the term born with CHD: 0.026 + 0.0088 = 0.0348.

#### Demographic parameters ($\varepsilon$, $\mu$)

We let individuals age out of their age group *i* and into the next age group *i*+1 at a rate $\epsilon_i$ defined as 1/time spent in the age group (in days). To keep the size of each age group constant, we added or subtracted a mortality/immigration constant $\mu_i$ in each age group:

$$\mu_i = \frac{n_{i-1}}{\epsilon_{i-1}} - \frac{n_i}{\epsilon_i}$$

#### Seasonally varying transmission rate ($\beta_{eff}$)

A seasonally varying transmission rate in directly transmitted diseases is commonly caused by a seasonal variation in contacts, susceptibility and pathogen transmissibility. For simplicity, we assumed that the effective transmission rate at time *t* ($\beta_{eff}$) followed a cosine function with an average value $\beta$, an amplitude $\eta$ and a phase shift $\varphi$:

$$\beta_{eff} = \beta * \left(1 + \eta * \cos\left(\frac{2\pi(t - 364\phi)}{365}\right)\right)$$

We assumed that the seasonality of contacts correlated with temperature, and that the maximum of contacts (i.e. the day when the seasonal forcing is maximal) occurred when temperature was at its lowest [17]. To determine the position of the maximal transmission rate (i.e the phase shift ϕ), we used the daily temperature data of eight randomly selected weather stations in Germany[1] and fit a sine curve to determine the day of the average temperature minimum.

[1] Downloaded from https://opendata.dwd.de/climate_environment/CDC/observations_germany/climate/daily/kl/historical/

#### Force of infection ($\lambda$)

The force of infection $\lambda$ is the rate at which susceptible individuals acquire the infection from an infected contact. In our model, λ varies by age group *i=1,…,25*, exposure level *l*=0,…,4, and over time through the seasonally varying transmission rate and the time-varying number of infectious individuals (symptomatic and asymptomatic). For each age-group, the force of infection depends on the number of contacts (susceptible) individuals from age-group *i* make with each age group *j* (given by the contact matrix *c*) and the share of currently infectious individuals in age group *j*. The infectious pressure from asymptomatically infected (*A*) is reduced by a factor $\alpha \in [0,1)$ compared to symptomatic infected (*I*). For each cycle of exposure and infection (reinfection level), the force of infection is reduced by a factor $\delta_l \in [0,1)$. The force of infection is therefore given by:

$$\lambda_i^l = \beta_{eff} * \delta_l * \sum_{j=1}^{25} \frac{\left(\alpha \sum_{k=0}^{4} A_j^k + \sum_{k=0}^{4} I_j^k\right) * c_{ij}}{n_j}$$

#### Contact matrix (c)

For the contact matrix we used data from the POLYMOD study [18]. Since the POLYMOD does not have contact data for monthly age groups, we used the available data for 0-1 year olds, divided the number of contacts by 12 and distributed them equally among the 12 monthly age groups. The contacts were then rescaled to make them reciprocal such that the total absolute number of contacts made from age group *i* to age group *j* corresponds to the total absolute number of contacts made from age group *j* to age group *i* (see script 1_prep_pop.R). The resulting contact matrix c is visualized in Fig S1, the numerical values are given in the table contacts_25_raw.csv in the GitHub repo.

Fig. S1. Contact matrix for the 25 age groups in the model. The contact patterns were informed by the POLYMOD study [18].

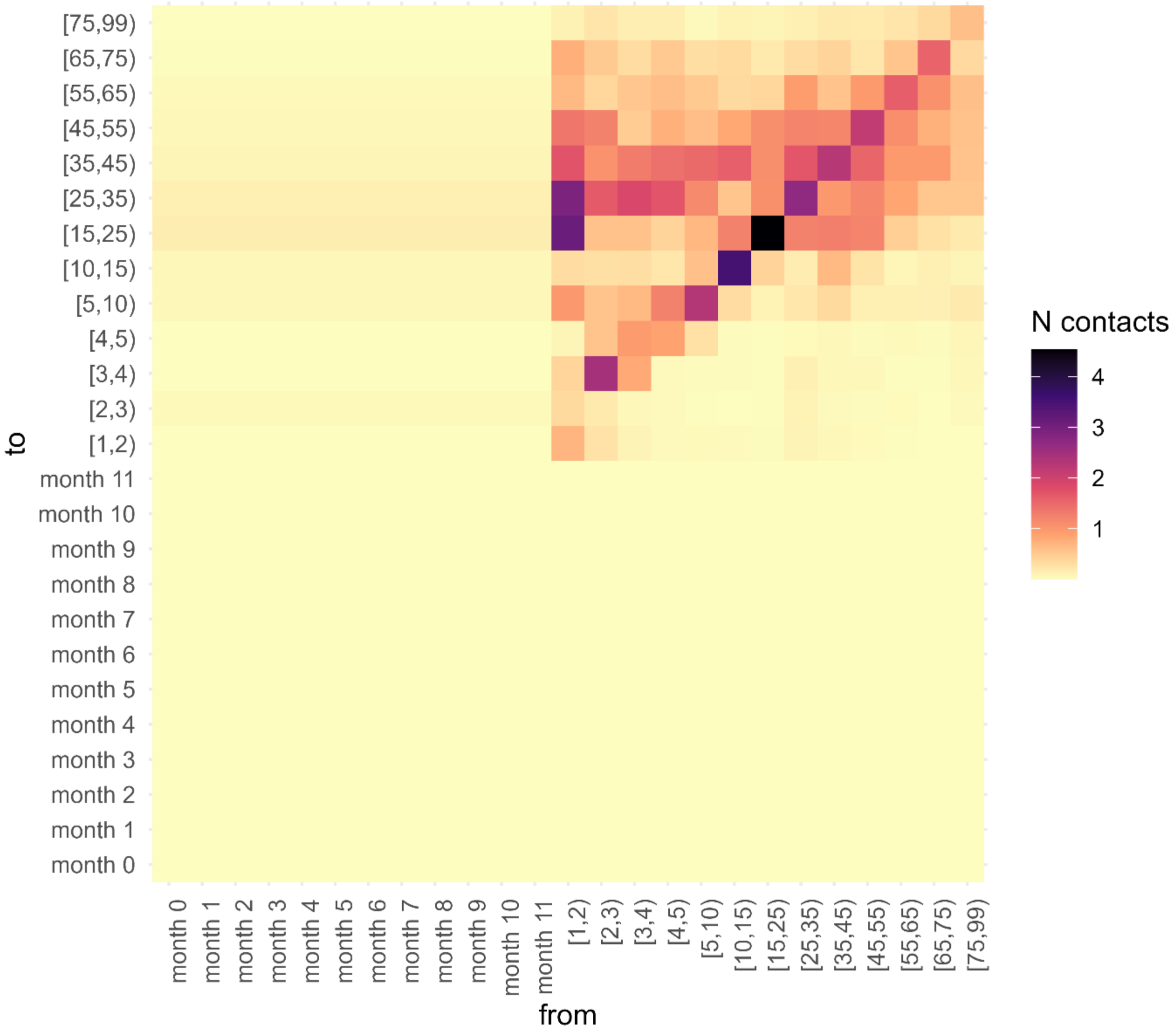

#### Duration of infectiousness (γ)

The duration of infectiousness in individuals who are infected for the first time was set to 9 days, corresponding to the average duration in 0-2 year olds measured by culture in a prospective cohort [5]. For simplicity, we assumed that the reduction of the duration of infectiousness was proportional by a factor *g* with each re-infection. Assuming that the duration in level 4 was 3.6 days [3], the proportional reduction factor *g* was calculated as 0.74 (i.e. 9 days * 0.74^3 = 3.6). Hence, the duration of infectiousness at level 2 is 6.7 days, the duration at level 3 is 4.9 days and the duration at level 4 is 3.6 days.

#### Calculation of age-dependent proportion of symptomatic cases detected (*q*)

For the calculation of the age-specific detection probability of symptomatic outpatient cases, *q*, we first calculated an annual detection probability $q^{annual}$, which was calculated as the ratio of the observed annual number of symptomatic cases (=detected in the AGI surveillance system, $C^{obs}$) and the average expected annual number of cases ($C^{exp}$) for each age group *i* for each year with data.

$$q_i^{annual} = \frac{C_i^{obs}}{C_i^{exp}}$$

The average expected annual number of cases by age group, $C^{exp}$, was calculated as the product of the age-specific attack rate (ar), the probability of symptomatic infection (ps) and the number of individuals in age group i (n):

$$C_i^{exp} = ar_i * ps_i * n_i$$

For the calculation of the attack rates and the proportion symptomatic see below. From the annual estimates of $q^{annual}$ we calculated a mean *q* for each age group (Fig S2A). Assuming that the true detection probability *q* lies somewhere between +/- 50% of the mean *q*, we defined a parameter $\rho_1$ with $\rho_1$ = [0.5 1.5], which is multiplied by the mean *q* to obtain the effective proportion of symptomatic cases detected in each age group *i* (Fig. S2B):

$$q_i = q_i^{mean} * \rho_1$$

The scaling parameter $\rho_1$ was fitted with a truncated Normal distribution with a mean of 1.0 and a standard deviation of 0.2 (see table S10). For simplicity, we assumed it to be constant across all age groups.

Fig S2. Observed (AGI data) and expected annual number of cases by age group (A). Annual ratio of observed and expected cases (black dots), mean detection probability $q^{mean}$ (red line) and upper (blue) and lower (green) assumed boundaries of the true detection probability *q* by age group (B).

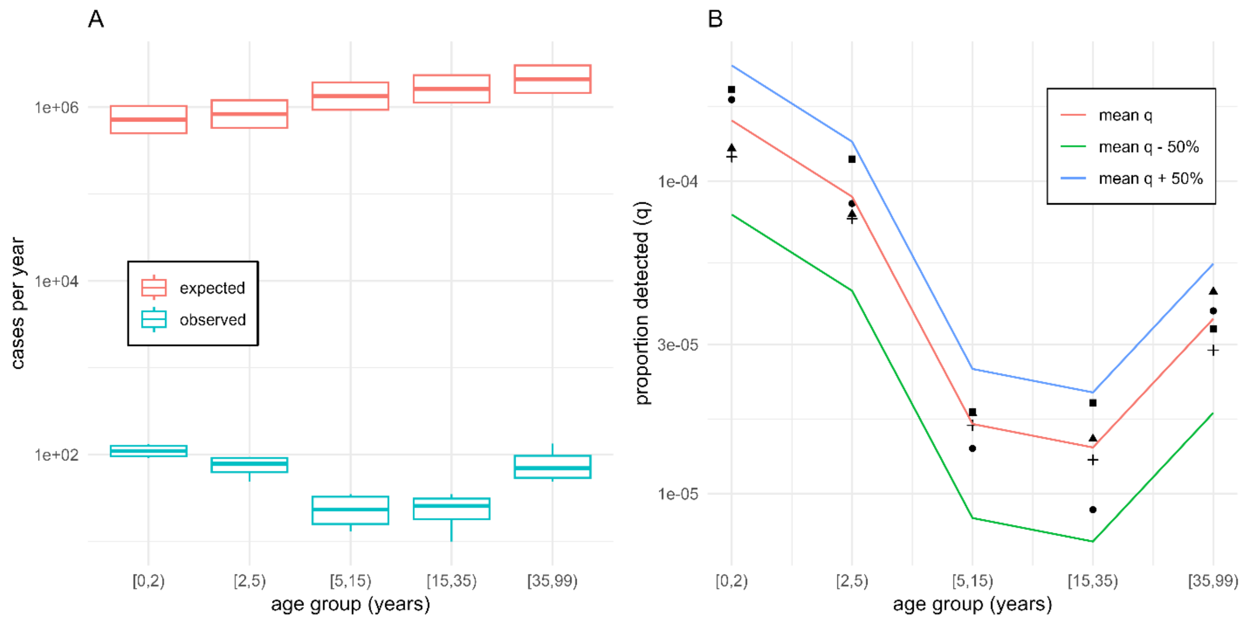

#### Calculation of age-dependent attack rates

To estimate the age-dependent attack rates (AR), we combined data from several studies and estimated an average age-dependent attack rate with a log-linear model from the mid-points of the age groups in the literature data (Fig S3A):

$$AR \sim log(age)$$

For the age groups <1 year old, we assumed an attack rate of 50%. We then predicted the age-specific attack rates for the age groups in the model (Fig S3B).

Fig. S3. Data and fitted age-specific annual attack rates (A(). Interpolated age-specific attack rates for all 25 age groups in the model (B).

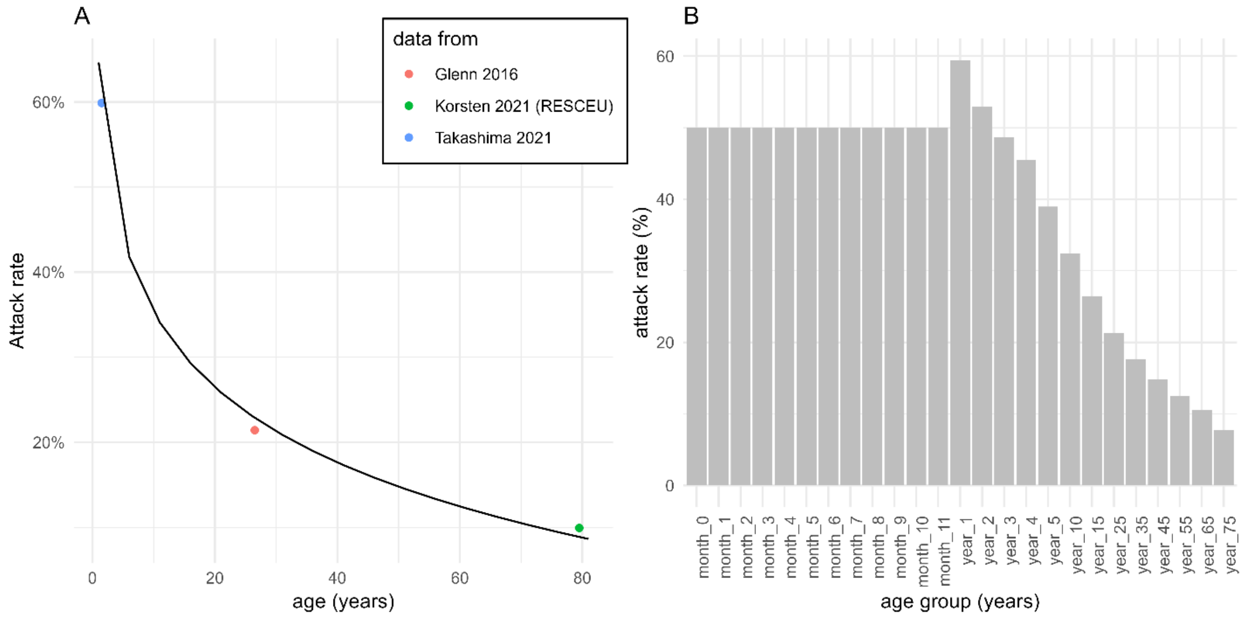

#### Calculation of age-dependent proportion of symptomatic cases reported ($p_s$)

To estimate the age-dependent probability of symptomatic disease, we used data from two studies: an observational household cohort from Kenya [4] for 0-64 year olds and an experimental challenge study [19] for 65+ year olds. Both studies tested individuals on a regular basis regardless of symptoms, and established the number of asymptomatic among all infected cases.

To interpolate the probabilities for each age group in the model, we fitted a GAM model with k=5 knots and a cubic regression splines smoothing term to the age group midpoints of the available data (Fig. SA). The proportion symptomatic (ps) were then estimated as 1-predicted proportion asymptomatic (pa) for each age group in the model (Fig. S3B).

Fig S4. Data and fitted age-specific probability of being asymptomatic when infected (black line) (A). Interpolated age-specific probabilities of being asymptomatic (B).

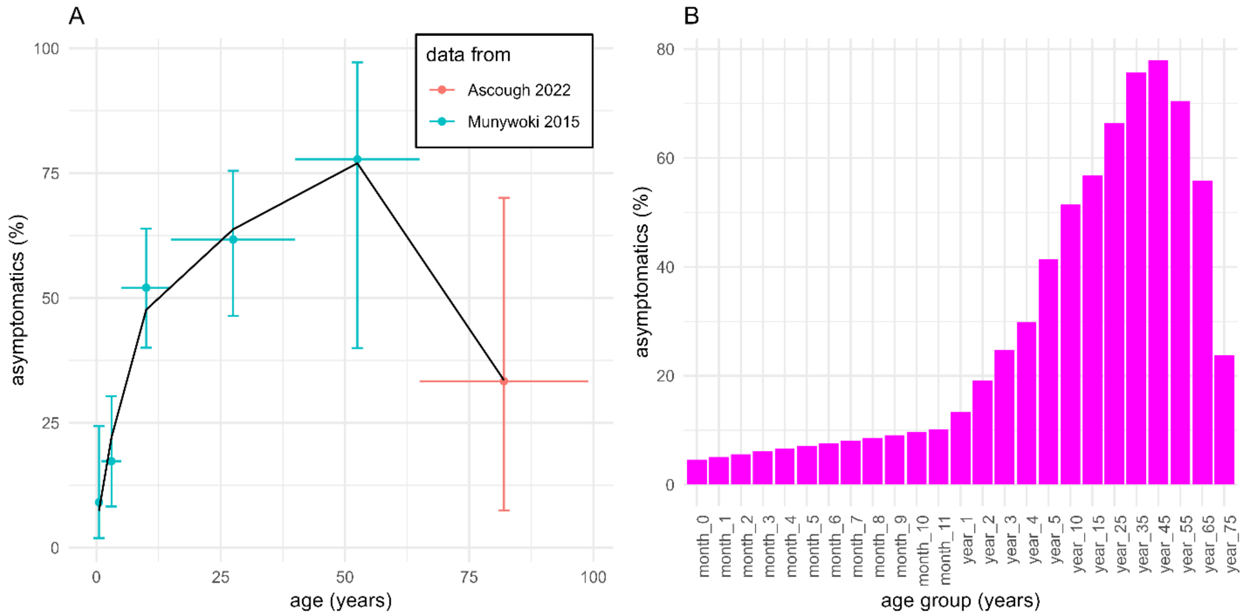

#### Calculation of age- and level-dependent proportion of symptomatic cases hospitalized (h)

For the calculation of the proportion hospitalised in 1+ year old, we again first calculated the annual expected number of symptomatic cases given the age-specific attack rate and the proportion symptomatic for all the years covered by the data (2015/2016-2018/2019) for each age group in the model (Fig S5A). We then divided the observed annual number of hospitalized cases by the expected annual number of symptomatic cases to obtain the proportion $h^{annual}$ for each year as a rough estimate for the age-specific probability of hospitalization over the four years covered in the data, which was then average over all years to obtain a mean *h* (Fig S5B). Assuming that the true *h* lies somewhere between +/- 50% of the mean *h*, we defined a parameter $\rho_2$ with $\rho_2$ = [0.5 1.5], which is multiplied by the mean *h* to obtain the effective proportion hospitalised in each age group *i*:

$$h_i = h_i^{mean} * \rho_2$$

The scaling parameter $\rho_2$ was fitted with a truncated Normal distribution with a mean of 1.0 and a standard deviation of 0.2 (see table S10). For simplicity, we assumed it to be constant across all age groups.

Fig S5. Observed and expected age-specific annual number of hospitalised cases (A). Annual ratios of observed and expected age-specific hospitalised cases (black dots), average ratio $q^{mean}$ (red line) of and lower (green line) and upper (blue line) assumed boundaries within which the true value of q would lie.

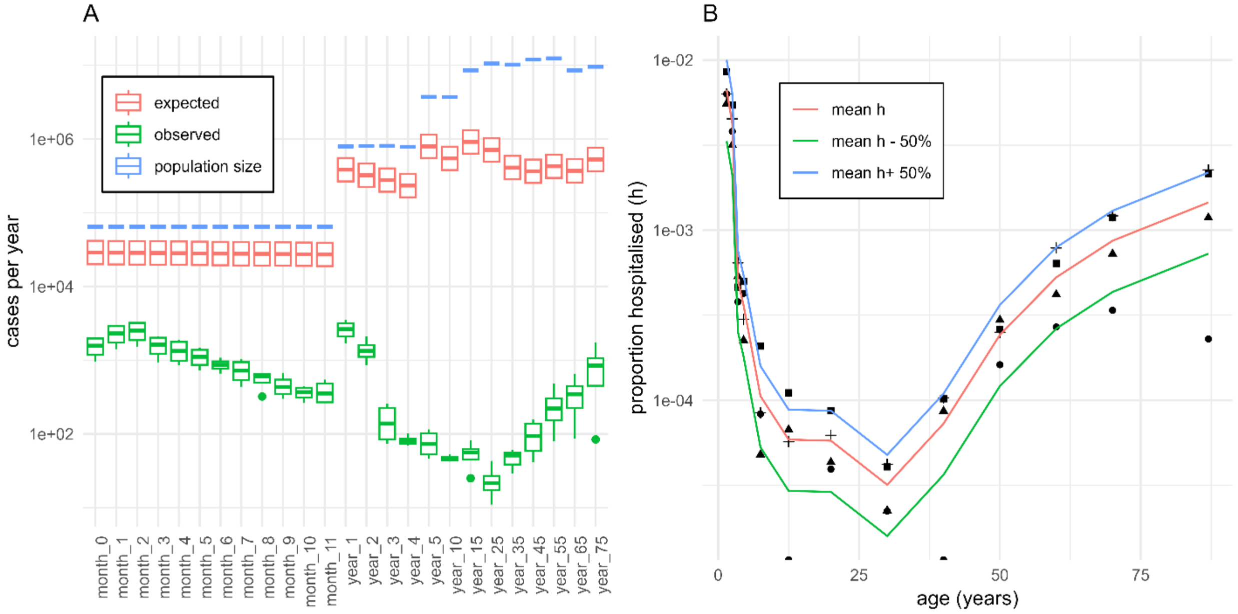

In the age groups <1 year old, the probability of hospitalization is much higher in the first few months after birth compared to the end of the first year, and we did not have sufficient data on the attack age-specific attack rates below 1 year of age. Therefore, we decided to fit this parameter entirely for <1 year old. We assumed that this probability was a function of age and followed an exponential decay with a lower bound at ages 11-12 months ($a_1$), and upper bound at ages 0-1 month ($a_2$) and a rate of decay ($a_3$) in both high and low risk infants. For simplicity, we modelled the probability of hospitalization in high risk infants, $h_h$, as

$$h_h = a_1 + a_2 * \exp(-a_3 * age)$$

All of the three parameters were fitted with uniform, bounded priors. The probability of hospitalization in low risk infants, $h_l$, was then calculated by scaling the probability of hospitalization in high risk infants ($h_h$) by 1/k, where k is the increased risk of hospitalization of high risk vs. low risk infants (see table with parameters).

$$h_l = h_h * \frac{1}{k}$$

#### Calculation of vaccine efficacy/protection based on published efficacy estimates from clinical trial data

Estimates on efficacy of immunisation products against medically attended symptomatic disease MAARI (hereafter denoted $VE_S$) and hospitalisations (hereafter denoted $VE_H$) from clinical trials are commonly an average (measured as a risk or rate ratio) over a specified observation time. However, these estimates are difficult to be used directly in a mathematical model, as the observed time window does not automatically correspond to the duration of protection, and the efficacy might decrease over time. To account for this, we simulated a separate waning model with an Erlang-2 distributed waning of protection. To estimate the initial vaccine efficacy estimates and the durations of protection adjusted for our model, we used published trial estimates of $VE_S$ and $VE_H$ and fitted the separate waning model with least-squares under the constraints that protection against hospitalisation is larger or equal to the protection against symptomatic disease and duration of protection against symptomatic disease is equal to the duration of protection against hospitalisation (code and data in Github repo script 3_prep_params.R). The resulting assumptions on protections for the different products and target groups used for the simulations are shown Figure S6. For Palivizumab, the vaccine efficacy is shown for an immunisation strategy of five monthly doses (one every 30 days).

Fig S6. Estimated decrease of vaccine efficacy over time based on fitted parameters for the initial vaccine efficacy for symptomatic medically-attended disease (top) and hospitalisation (bottom), and the duration of protection, for the different immunisation products (Palivizumab, Nirsevimab, maternal vaccine in the infant, maternal vaccine in the model, older adult vaccine). All immunisation products are modelled as a single dose except Palivizumab, which is modelled with 5 injections.

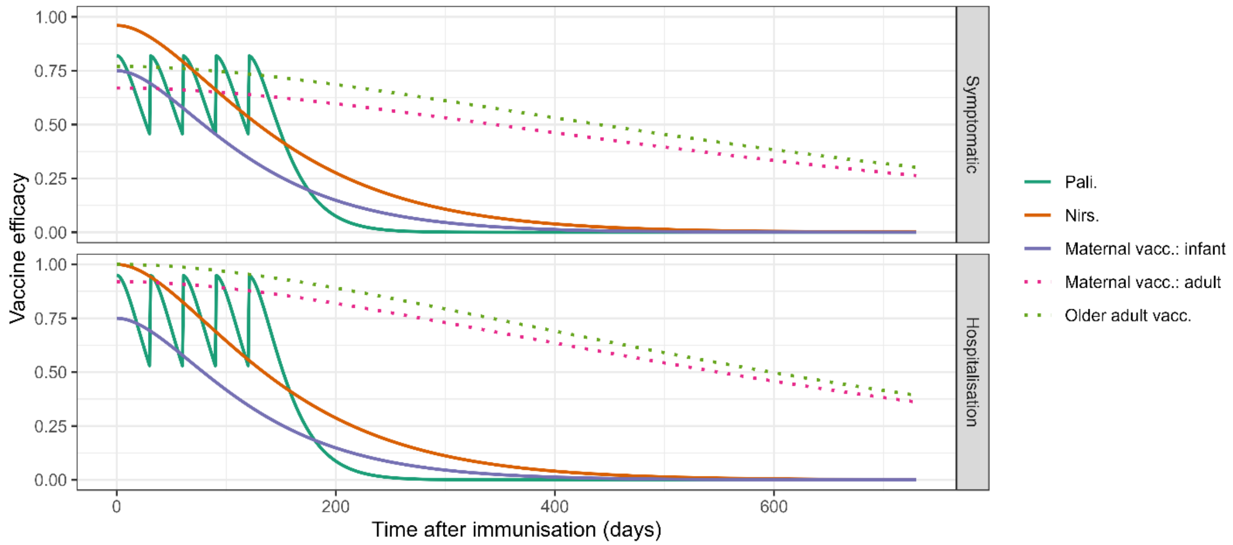

#### Calculation of model derived quantities

The outcomes of the vaccination simulation model were the weekly incident numbers of symptomatic RSV cases, total (symptomatic and asymptomatic) cases, RSV-related hospitalisations, ICU stays, and RSV-related (in-hospital) mortality. We calculated the annual incidences in each age group, and we estimated the absolute and relative number of cases averted by the alternative immunisation strategies as compared to the baseline scenario. We also present results on the efficiency in terms of the number needed to vaccinate (NNV) to prevent one RSV case, one RSV-related hospitalisation, one RSV-related ICU stay, and one RSV-related (in-hospital) death. As we assumed an identical number of immunized patients when switching immunisation in high-risk infants, we cannot provide NNV estimates for the endpoints in this scenario.

#### Estimation of age-specific hospitalization-ICU and hospitalization-fatality ratio

To estimate the risk of severe disease endpoints among hospitalised individuals (ICU and death), we used routinely collected hospitalisation data from the 'Institut fuer das Entgeltsystem im Krankenhaus' (InEK) based on primary diagnosis codes of all yearly RSV-specific admissions to all hospitals in Germany in 2019-2023 (pre-pandemic data were not available). We estimated age-group specific hospitalisation-ICU and hospitalisation-mortality rates (assumed as time-constant over the five years) based on a generalized linear model with Poisson-link using the yearly ICU or mortality counts per age-group as outcome and the corresponding (log-) number of hospitalisations as offset. These rates were then used to calculate the proportions of hospitalised patients who would be referred to ICU or who would die.

The resulting ratios were applied as multiplication/scaling factors to the simulated yearly hospitalisation counts from the dynamic transmission model when simulating disease burden for different strategies of RSV immunisation. Uncertainty was propagated via the different analysis steps by sampling one set of age-specific hospitalisation-ICU and hospitalisation-mortality rates per posterior draw of the parameters from the calibrated dynamic transmission model used for simulation.

Fig. S7. Estimated probabilities of ICU referral (red) and mortality (blue) when hospitalised, by age group based on RSV-specific InEK data. Shown are point estimates and 95% prediction intervals.

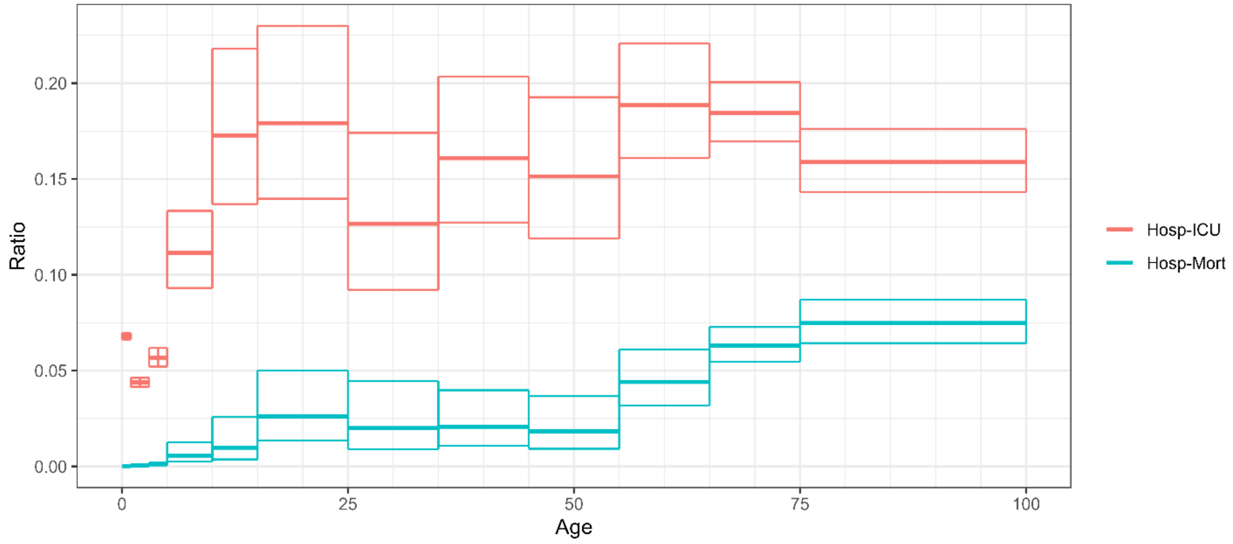

#### Estimation of under-ascertainment of RSV-related hospitalisations in older adults

RSV is not routinely diagnosed in hospitalised older adults with acute respiratory infections. Therefore, hospitalisation data based on RSV-specific ICD-10 codes are likely an underrepresentation of the true burden of disease. We therefore estimated a factor to scale the simulated hospitalisations in older adults up to represent the likely burden. To calculate the scaling factor, we compared the model derived hospitalisation rates to estimates of RSV-related hospitalisations in older adults from systematic reviews or statistical modelling of population based studies for Germany and/or industrialized or high-income [20–22] countries.

### Results

#### Fitted model

Fig S8. Age-specific quarterly hospitalisations (A) and weekly outpatient visits (B) simulated from the model (red bars/lines) and compared to the data (black dots).

A

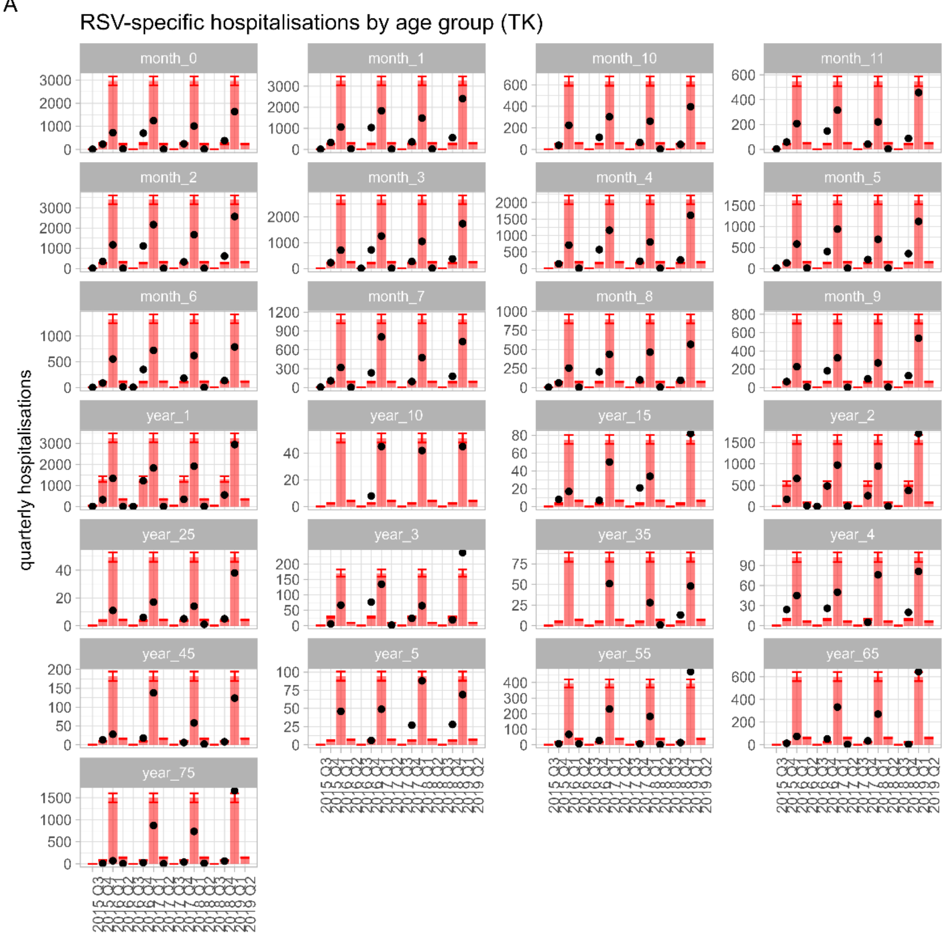

B

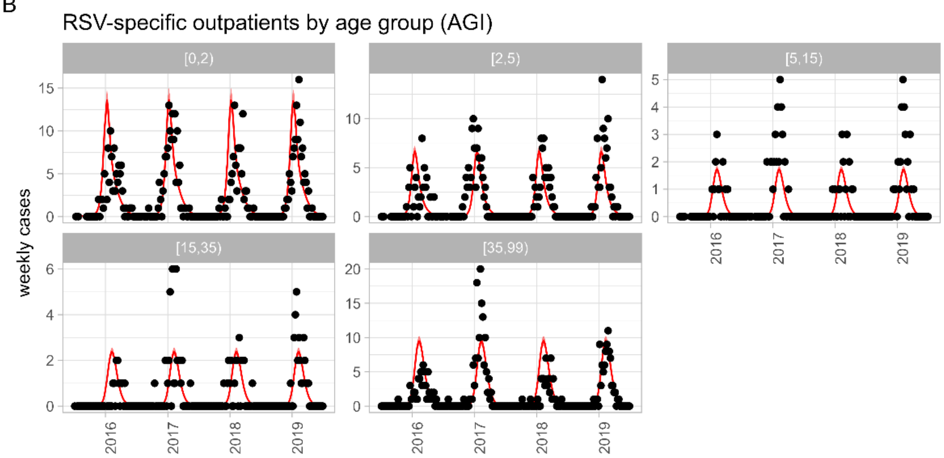

Fig. S9. Trace plots of fitted model.

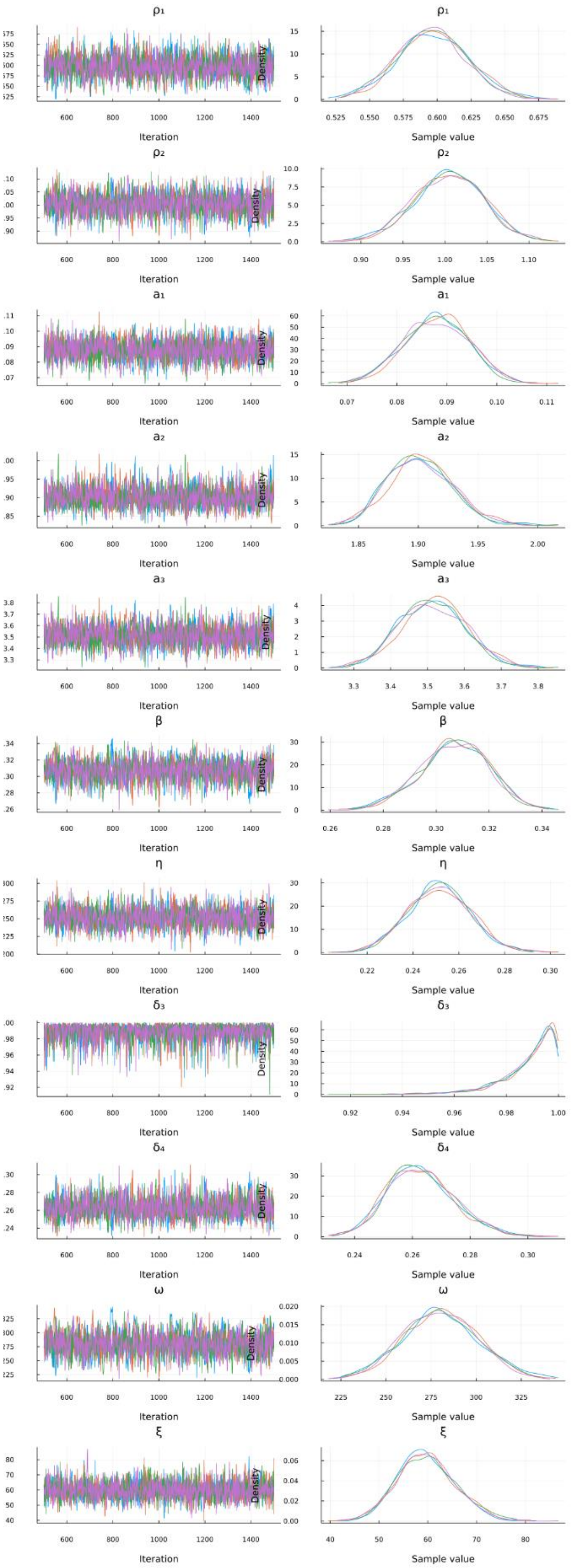

Table S13: Marginal posterior estimates (median, 95% CrI).

| parameter | 2.50% | 50.00% | 97.50% |
|---|---|---|---|
| $\rho_1$ | 0.545587 | 0.596181 | 0.648259 |
| $\rho_2$ | 0.921904 | 1.005193 | 1.083711 |
| $a_1$ | 0.075046 | 0.088011 | 0.100129 |
| $a_2$ | 1.852254 | 1.900292 | 1.959913 |
| $a_3$ | 3.341473 | 3.513478 | 3.696081 |
| β | 0.280375 | 0.307137 | 0.330915 |
| η | 0.225292 | 0.251413 | 0.27871 |
| $\delta_3$ | 0.960213 | 0.992276 | 0.999769 |
| $\delta_4$ | 0.24214 | 0.262241 | 0.287655 |
| ω | 239.8174 | 279.9774 | 321.7776 |
| ξ | 48.84686 | 59.50121 | 71.9559 |

Table S14. Annual number of RSV cases by age group based on the model prediction for the current state (Palivizumab only for high risk infants). Estimates are medians [95% posterior predictive intervals]. The hospitalisations are not adjusted for under-ascertainment, hence they represent the RSV-specific hospitalisations capture with the four RSV-specific ICD-10 codes.

| age group | infected cases | symptomatic cases | hospitalised | icu | deaths |
|---|---:|---:|---:|---:|---:|
| 1 | 10,038 [9,547, 10,558] | 9,593 [9,124, 10,090] | 3,444 [3,275, 3,621] | 234 [223, 247] | 1 [0, 1] |
| 2 | 11,913 [11,328, 12,451] | 11,084 [10,541, 11,585] | 3,786 [3,602, 3,957] | 258 [242, 273] | 1 [0, 1] |
| 3 | 12,560 [11,955, 13,081] | 11,624 [11,063, 12,104] | 3,966 [3,776, 4,129] | 269 [254, 282] | 1 [0, 1] |
| 4 | 12,890 [12,268, 13,425] | 11,878 [11,305, 12,369] | 3,127 [2,968, 3,253] | 212 [201, 223] | 1 [0, 1] |
| 5 | 13,051 [12,447, 13,623] | 11,967 [11,416, 12,492] | 2,441 [2,327, 2,550] | 166 [157, 175] | 0 [0, 1] |
| 6 | 13,133 [12,543, 13,733] | 11,985 [11,446, 12,530] | 1,919 [1,830, 2,010] | 130 [124, 136] | 0 [0, 0] |
| 7 | 13,161 [12,587, 13,783] | 12,051 [11,526, 12,619] | 1,563 [1,491, 1,642] | 106 [101, 112] | 0 [0, 0] |
| 8 | 13,162 [12,593, 13,786] | 12,059 [11,538, 12,630] | 1,277 [1,217, 1,342] | 87 [82, 92] | 0 [0, 0] |
| 9 | 13,137 [12,566, 13,754] | 12,010 [11,488, 12,574] | 1,050 [999, 1,105] | 72 [67, 75] | 0 [0, 0] |
| 10 | 13,080 [12,515, 13,695] | 11,913 [11,398, 12,473] | 875 [829, 921] | 59 [56, 63] | 0 [0, 0] |
| 11 | 13,012 [12,450, 13,617] | 11,796 [11,286, 12,345] | 742 [699, 784] | 50 [47, 53] | 0 [0, 0] |
| 12 | 12,932 [12,377, 13,529] | 11,664 [11,164, 12,202] | 641 [601, 680] | 44 [41, 47] | 0 [0, 0] |
| 13 | 862,145 [822,626, 900,308] | 747,026 [712,783, 780,093] | 4,922 [4,640, 5,175] | 216 [197, 232] | 3 [2, 4] |
| 14 | 655,681 [610,806, 701,110] | 529,851 [493,589, 566,562] | 2,224 [2,096, 2,340] | 98 [90, 104] | 1 [1, 2] |
| 15 | 525,584 [494,544, 556,617] | 395,636 [372,270, 418,995] | 209 [196, 220] | 12 [10, 13] | 0 [0, 0] |
| 16 | 427,033 [406,948, 448,227] | 299,775 [285,675, 314,652] | 118 [111, 124] | 7 [6, 7] | 0 [0, 0] |
| 17 | 1,723,166 [1,636,585, 1,823,866] | 1,008,707 [958,024, 1,067,655] | 108 [102, 113] | 12 [10, 14] | 1 [0, 1] |
| 18 | 1,667,547 [1,566,732, 1,784,054] | 809,390 [760,457, 865,940] | 58 [55, 61] | 10 [8, 13] | 1 [0, 1] |
| 19 | 3,617,913 [3,389,558, 3,881,397] | 1,559,915 [1,461,456, 1,673,520] | 85 [81, 90] | 15 [13, 20] | 2 [1, 4] |
| 20 | 4,647,999 [4,368,549, 4,967,220] | 1,562,744 [1,468,787, 1,670,072] | 57 [54, 60] | 7 [5, 10] | 1 [0, 2] |
| 21 | 4,020,013 [3,787,186, 4,290,516] | 976,031 [919,502, 1,041,707] | 95 [90, 100] | 15 [12, 19] | 2 [1, 4] |
| 22 | 4,352,145 [4,109,692, 4,648,336] | 956,109 [902,846, 1,021,178] | 209 [197, 219] | 31 [25, 40] | 4 [2, 7] |
| 23 | 3,431,614 [3,249,981, 3,659,289] | 1,012,919 [959,306, 1,080,123] | 451 [427, 474] | 85 [73, 95] | 20 [14, 27] |
| 24 | 2,184,263 [2,067,684, 2,323,548] | 963,891 [912,445, 1,025,355] | 688 [652, 725] | 126 [116, 139] | 44 [37, 51] |
| 25 | 2,053,462 [1,952,785, 2,170,141] | 1,563,871 [1,487,198, 1,652,731] | 1,720 [1,634, 1,817] | 272 [249, 315] | 130 [111, 153] |
| total | 30,315,668 [28,664,984, 32,261,791] | 12,529,517 [11,863,475, 13,298,194] | 35,789 [33,933, 37,363] | 2,600 [2,467, 2,721] | 213 [189, 244] |

Figure S10. Fitted probability of a symptomatic case being hospitalised and reported as RSV-specific ICD10 codes in high and low risk infants (A) and low risk individuals 1+ year olds (B).

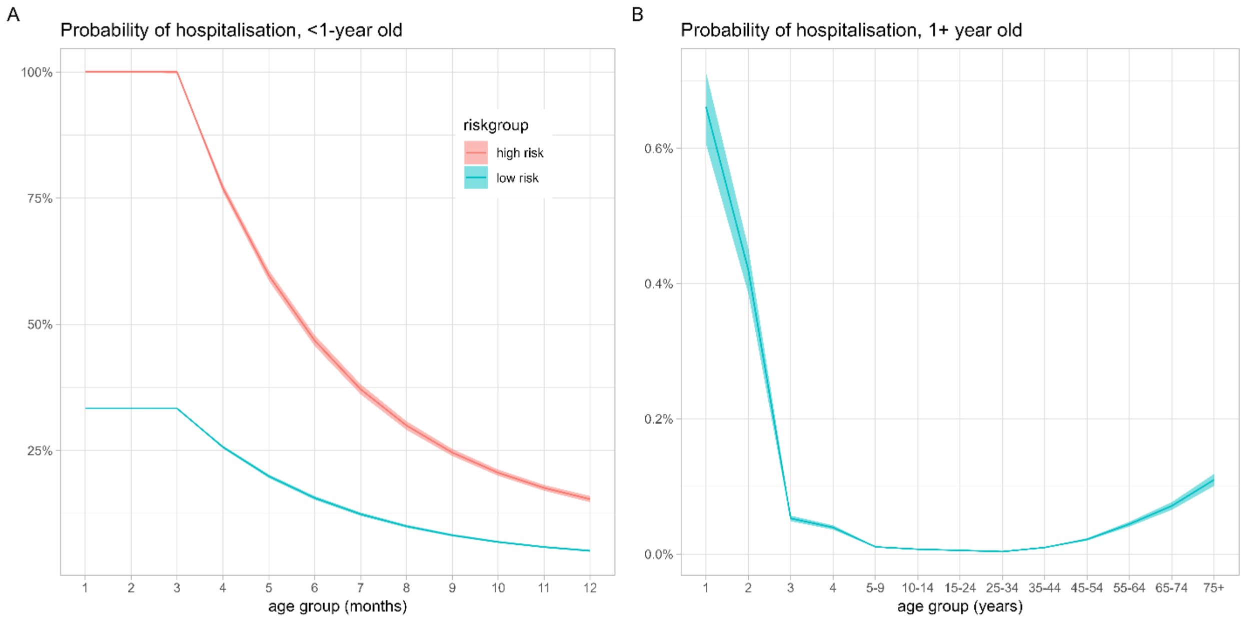

#### Quantification of under-ascertainment in RSV-related hospitalization data for older adults

While the calibrated model represents RSV-specific ICD10-code derived hospitalizations from InEK well, it is obvious, that the model-based hospitalization incidence in older adults is, depending on the specific age-group and study, reduced by factor 8-14 compared to the systematic reviews on RSV-related hospitalisation incidence. We therefore scaled up the simulation-based hospitalisation counts as well as derived quantities (ICU numbers and in-hospital deaths) by a factor 8-14 for our main analysis of the effects of older adult vaccination strategies and performed a sensitivity analysis on the effect of this upscaling for under-ascertainment factors from 1 (no under-ascertainment) to 15.

Fig. S11. Model-based, yearlyRSV hospitalisation rates after calibration (blue lines) and literature-derived rates (brown, yellow and green lines) for 60+ year olds by age group. The ratio of these two estimates yields the scaling factor for the under-ascertainment. Grey bars show reported hospitalisation rates from InEK.

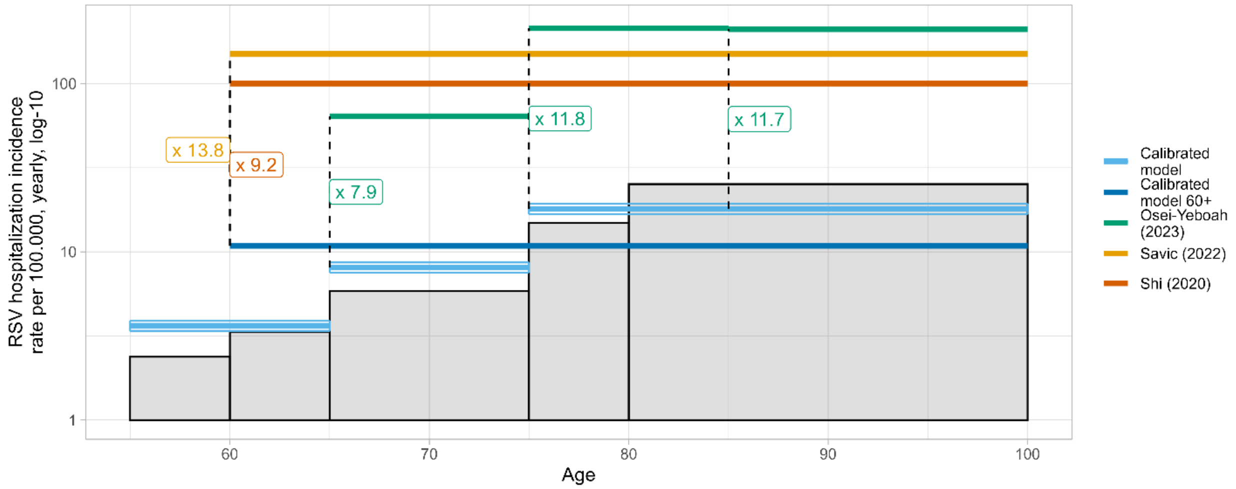

#### Vaccination simulations

Fig S12: **Model-based incidence of symptomatic RSV and severe disease endpoints per age group and simulation year at the infant base strategy 0 (immunisation of high-risk infants with Palivizumab)**. Panel A shows the annual absolute number of expected cases in each age-group per simulated endpoint (rows) and simulation year (columns). Panel B shows the corresponding rate per 100.000 individuals.

A

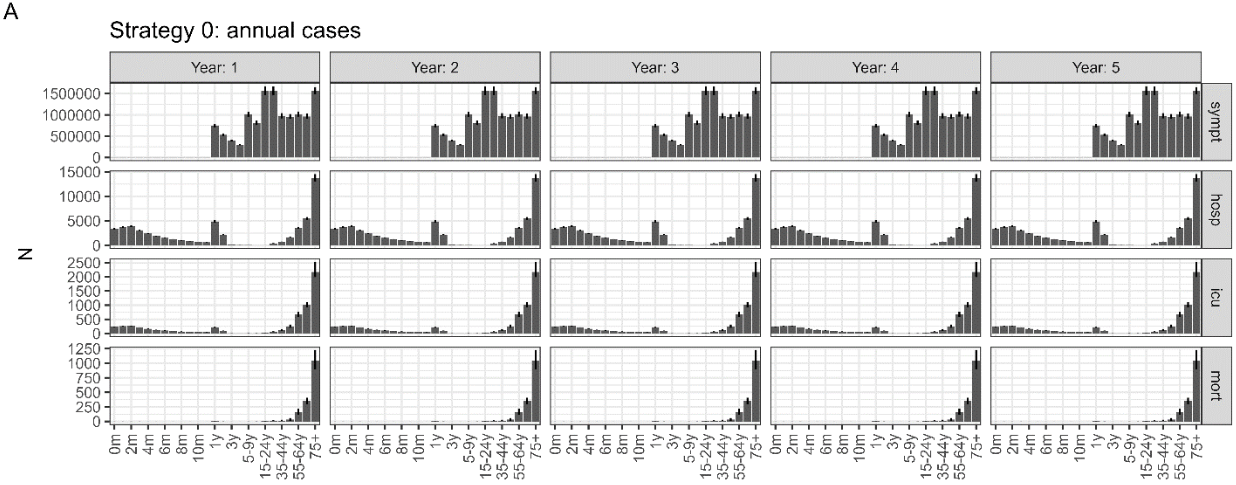

B

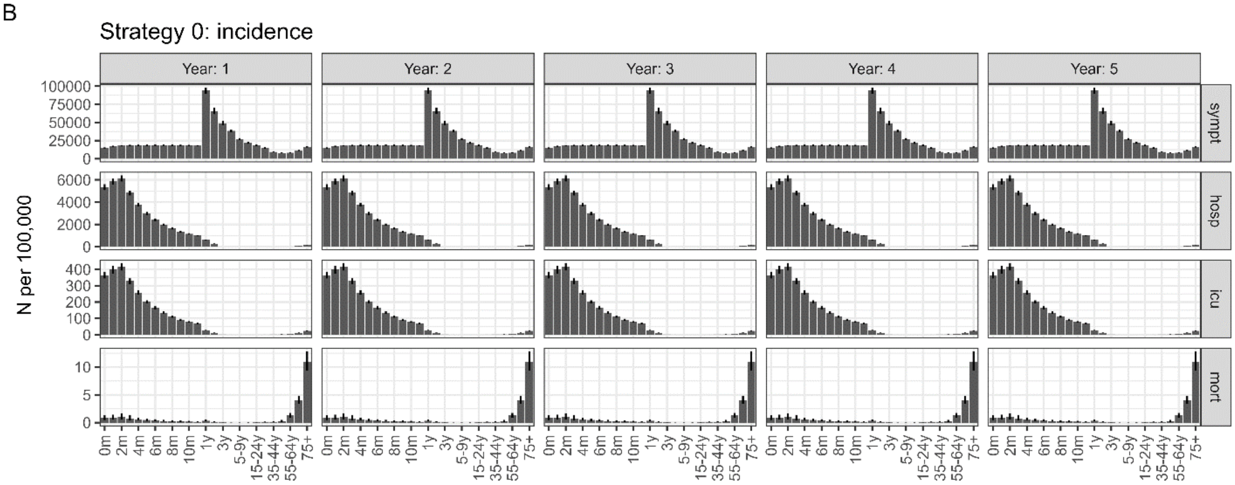

Fig S13: **Model-based prevented incidence of symptomatic RSV and severe disease endpoints per age-group and simulation year for immunisation strategy 1 (long-acting mABs in high-risk infants) compared to the infant baseline scenario**. Panel A shows the expected absolute number of prevented cases for each age-group per simulated endpoint (rows) and simulation year (columns). Panel B shows the corresponding rate per 100.000 individuals. Panel C shows the relative reduction in case numbers compared to the baseline scenario.

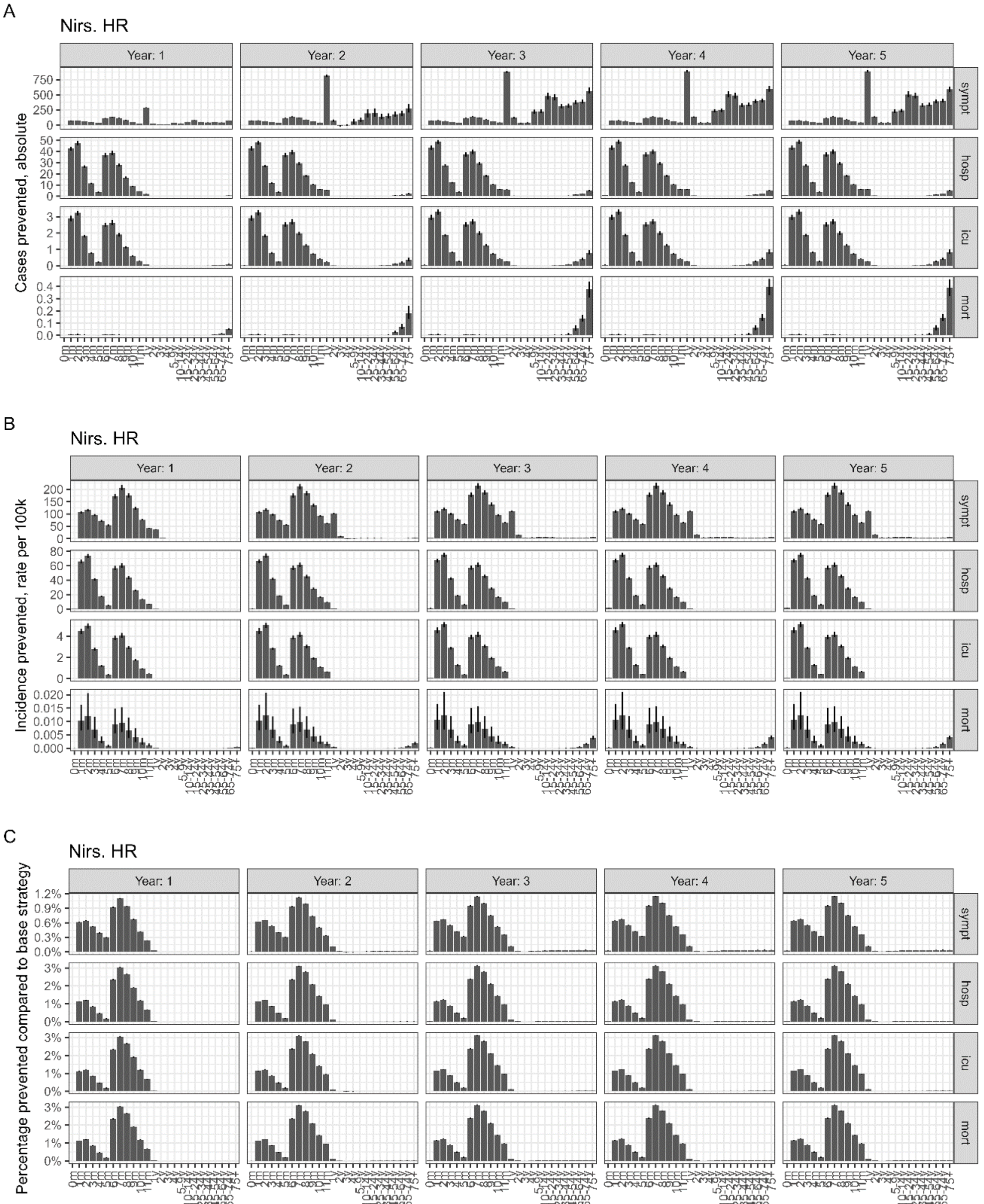

Fig S14: **Model-based prevented incidence of symptomatic RSV and severe disease endpoints per age-group and simulation year for immunisation strategy 2 (long-acting mABs in all infants) compared to the infant baseline scenario**. Panel A shows the expected absolute number of prevented cases for each age-group per simulated endpoint (rows) and simulation year (columns). Panel B shows the corresponding rate per 100.000 individuals. Panel C shows the relative reduction in case numbers compared to the baseline scenario.

A

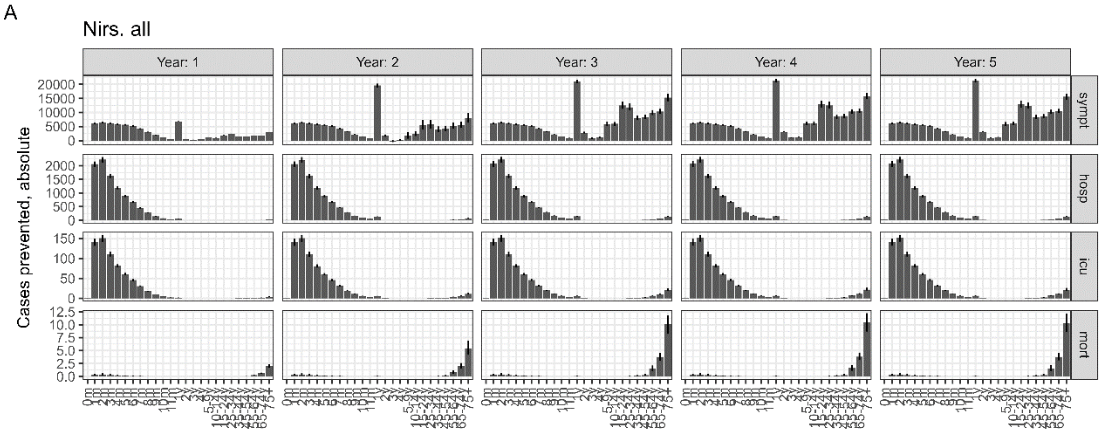

B

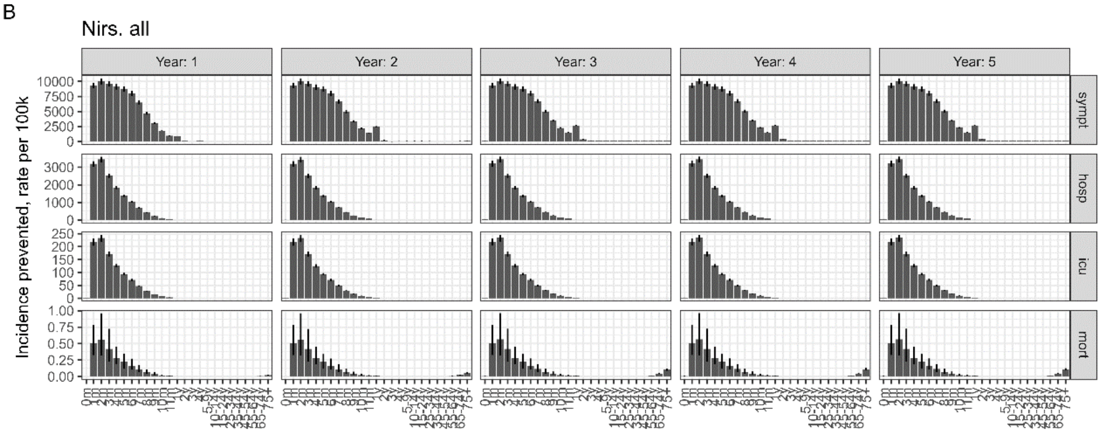

C

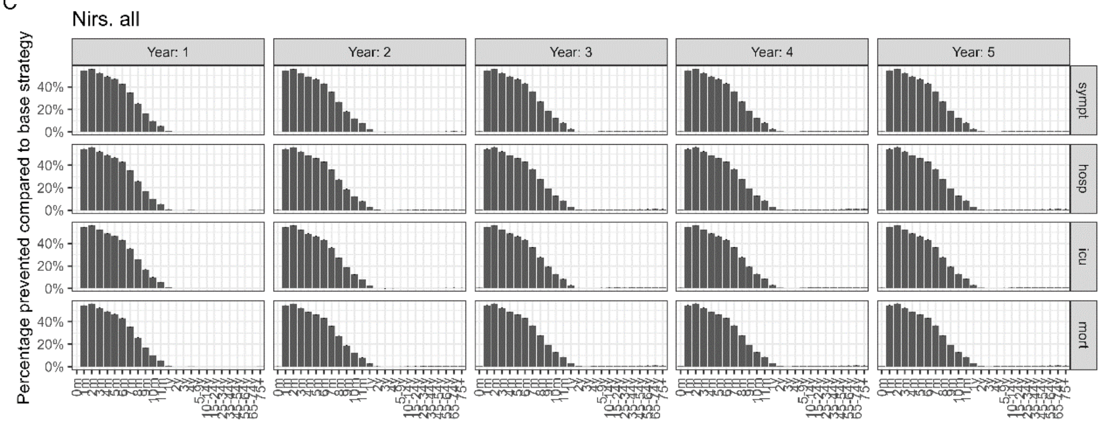

Fig S15: **Model-based prevented incidence of symptomatic RSV and severe disease endpoints per age-group and simulation year for immunisation strategy 3 (seasonal maternal vaccination and Palivizumab in high risk) compared to the infant baseline scenario (Palivizumab in high risk)**. Panel A shows the expected absolute number of prevented cases for each age-group per simulated endpoint (rows) and simulation year (columns). Panel B shows the corresponding rate per 100.000 individuals. Panel C shows the relative reduction in case numbers compared to the baseline scenario.

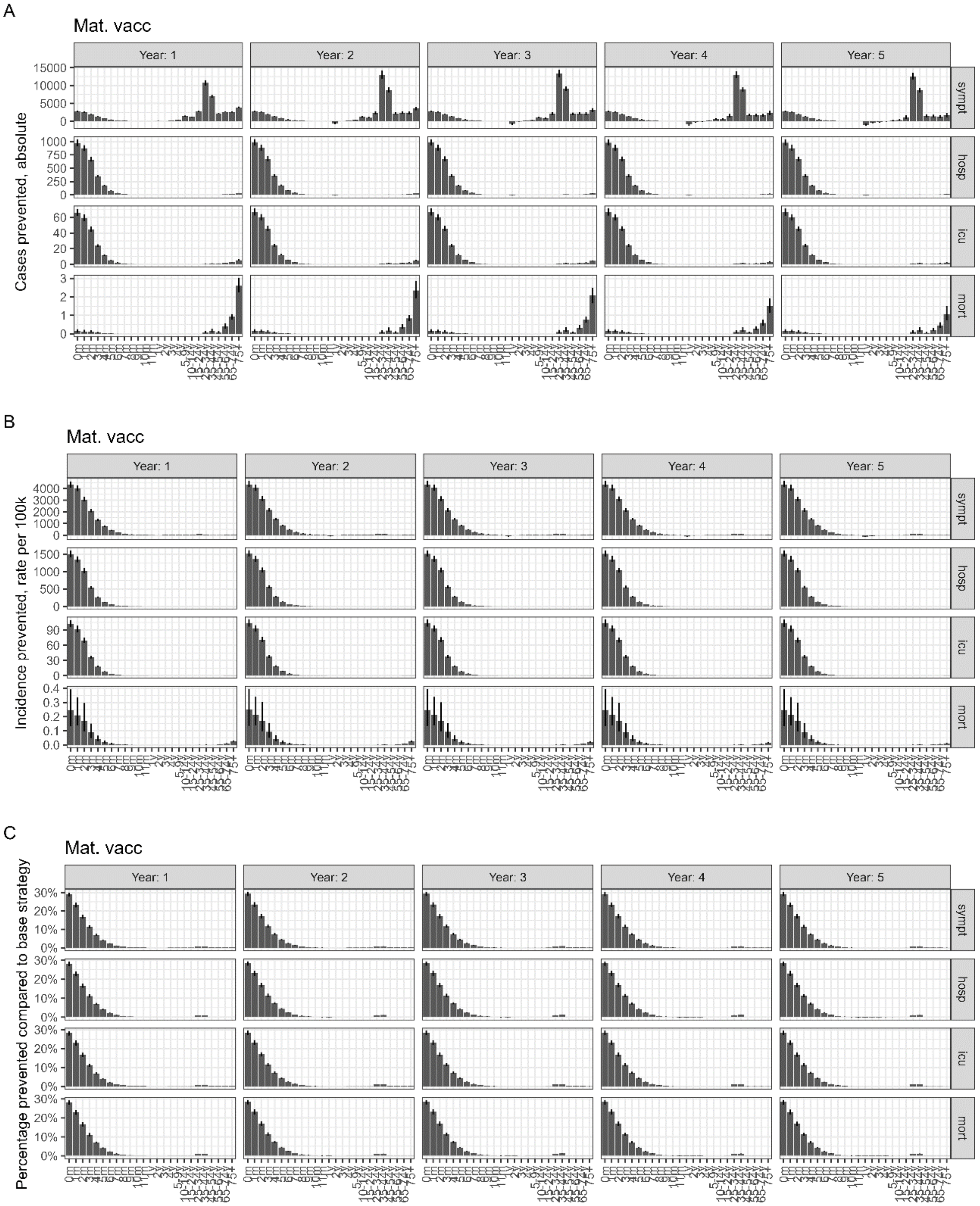

Fig S16: Share of prevented incidence per simulation year (among total prevented incidence over 5 years) for the different immunisation strategies. The strategies targeting infants show a constant reduction in hospitalisation, ICU hospitalisation and mortality over the five projected years. The prevention of symptomatic cases in the infant strategies increases over time due to an increasing indirect effect (reduction of symptomatic cases among immunised leading to lower onwards transmission and hence also fewer symptomatic cases in other age groups). The strategies in older adults on the other side prevent most cases in the first year, while the effect decreases in the following years due to waning of immunity.

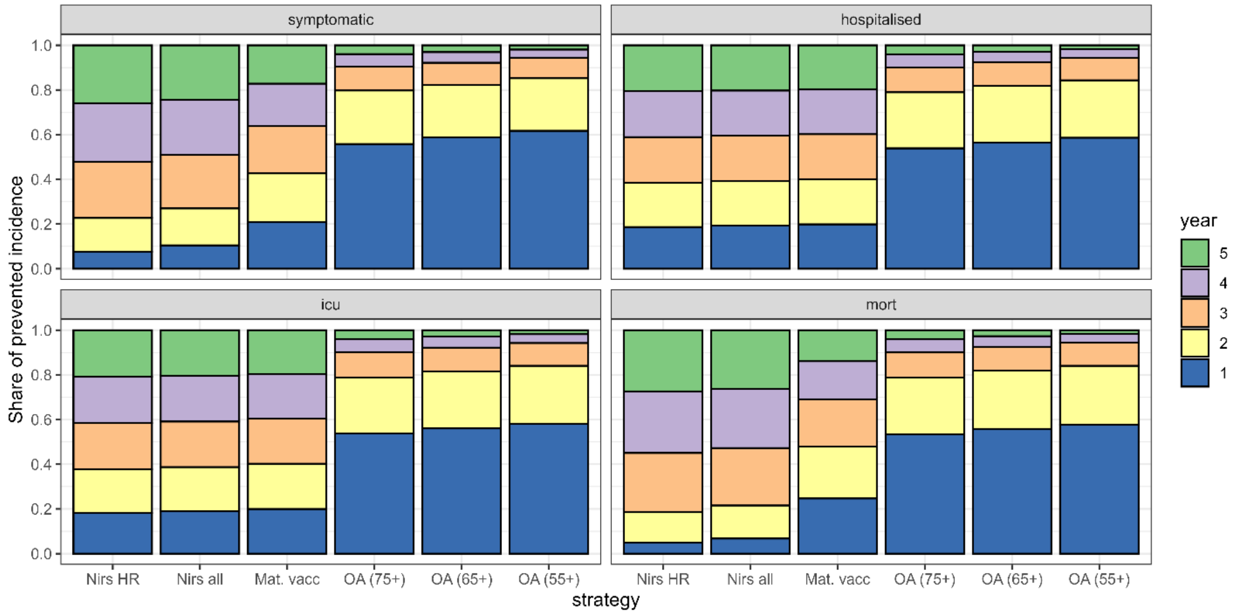

Figure S17: Sensitivity analysis for different uptake levels of (seasonal) infant RSV immunisation with either Nirsevimab in infants aged month 1-5 (purple) or a seasonal maternal vaccination programme and Palivizumab for high risk (green) (A), and the total number of immunised individuals in either infant RSV immunisation programme (B). The dashed lines connect the averted percentages of cases in both strategies (Nirsevimab and maternal) that correspond to the same modelled uptake in the target group.

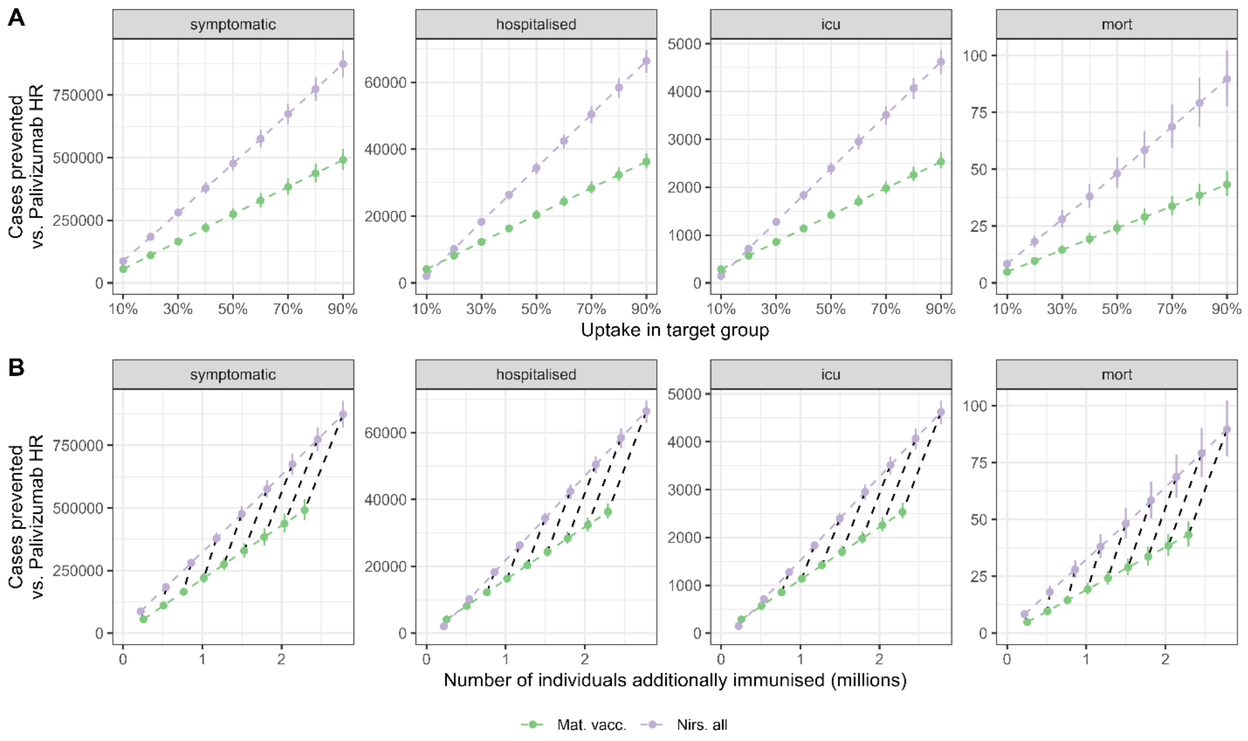

Fig. S18: Sensitivity analysis on the implementation of a seasonal vs. year-round maternal vaccination programme.

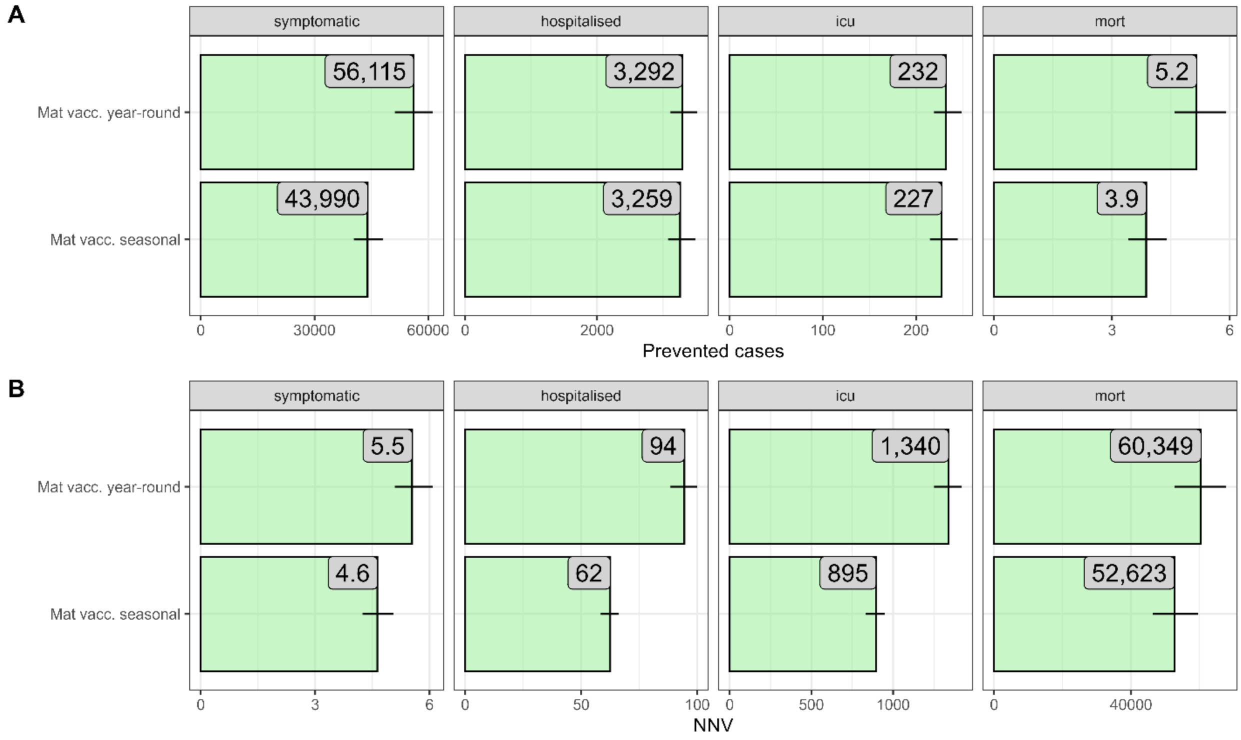

Fig. S19: **Model-based incidence of symptomatic RSV and severe disease endpoints per age-group and simulation year at the older adult baseline scenario (immunisation in all infants with Nirsevimab, 70% uptake)**. Panel A shows the absolute number of expected cases in each age-group per simulated endpoint (rows) and simulation year (columns). Panel B shows the corresponding rate per 100.000 individuals. Expected hospitalisation counts in adults (as well as derived ICU/mortality endpoints) are adjusted for under-ascertainment based on scaling factor 8.

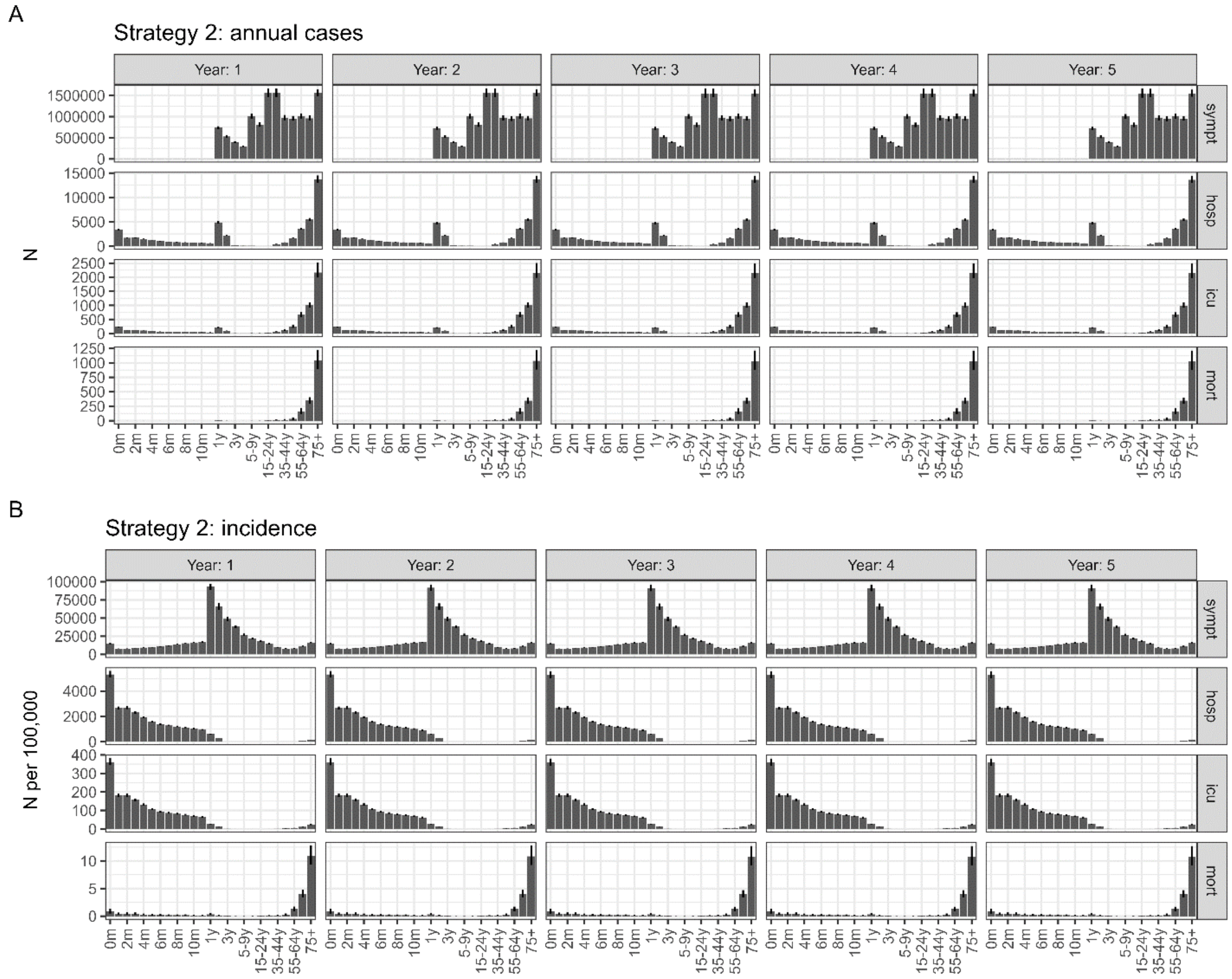

Fig S20: **Model-based prevented incidence of symptomatic RSV and severe disease endpoints per age-group and simulation year for immunisation scenario 5 (older adult vaccination) in 75+ compared to the older adult baseline scenario**. Panel A shows the expected absolute number of prevented cases for each age-group per simulated endpoint (rows) and simulation year (columns). Panel B shows the corresponding rate per 100.000 individuals. Panel C shows the relative reduction in case numbers compared to the baseline scenario. Expected hospitalisation counts in adults (as well as derived ICU/mortality endpoints) are adjusted for under-ascertainment based on scaling factor 8.

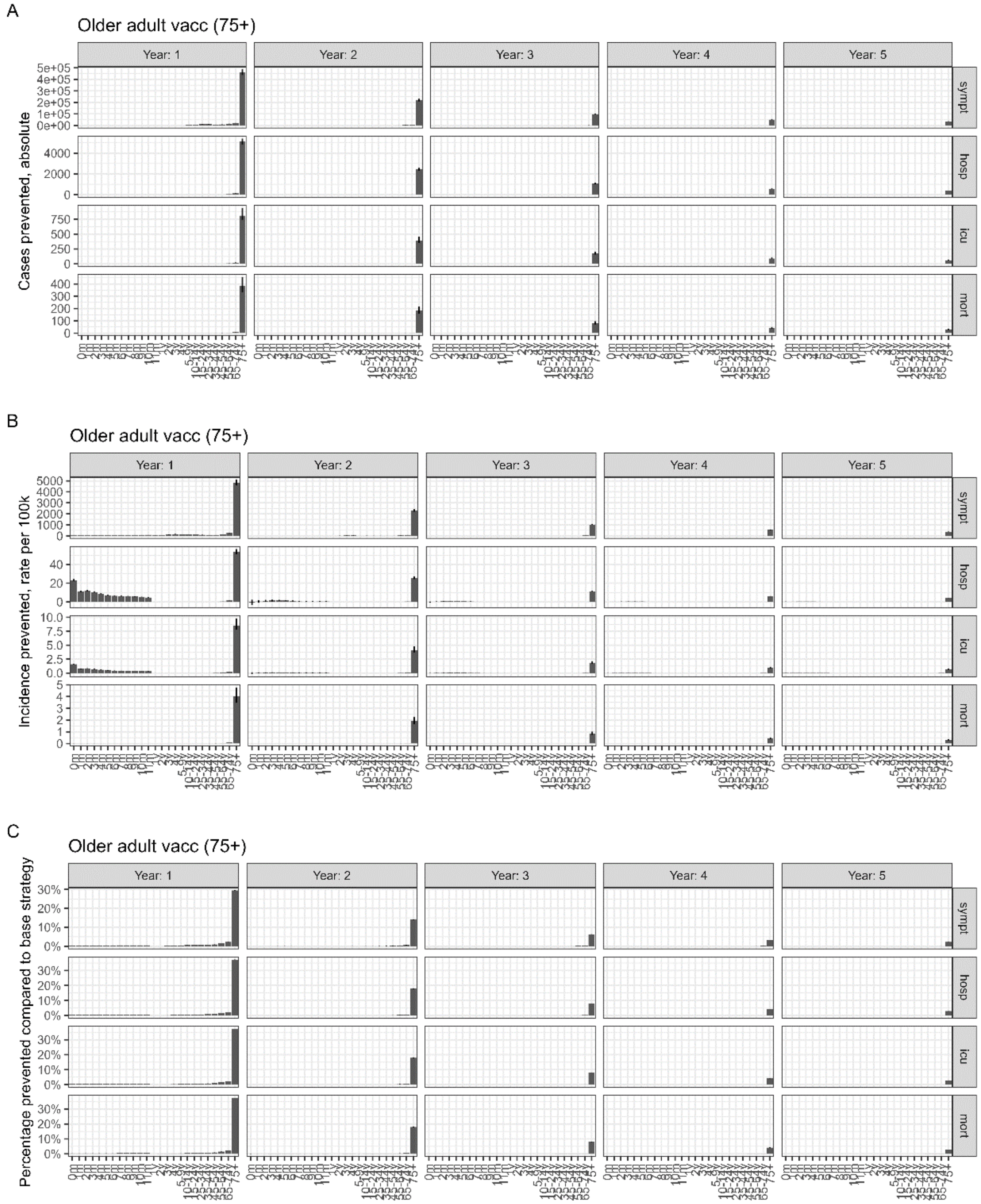

Fig S21: **Model-based prevented incidence of symptomatic RSV and severe disease endpoints per age-group and simulation year for immunisation scenario 5 (older adult vaccination) in 65+ compared to the older adult baseline scenario**. Panel A shows the expected absolute number of prevented cases for each age-group per simulated enpoint (rows) and simulation year (columns). Panel B shows the corresponding rate per 100.000 individuals (i.e., person-years per age-group). Panel C shows the relative reduction in case numbers compared to the baseline scenario.

A

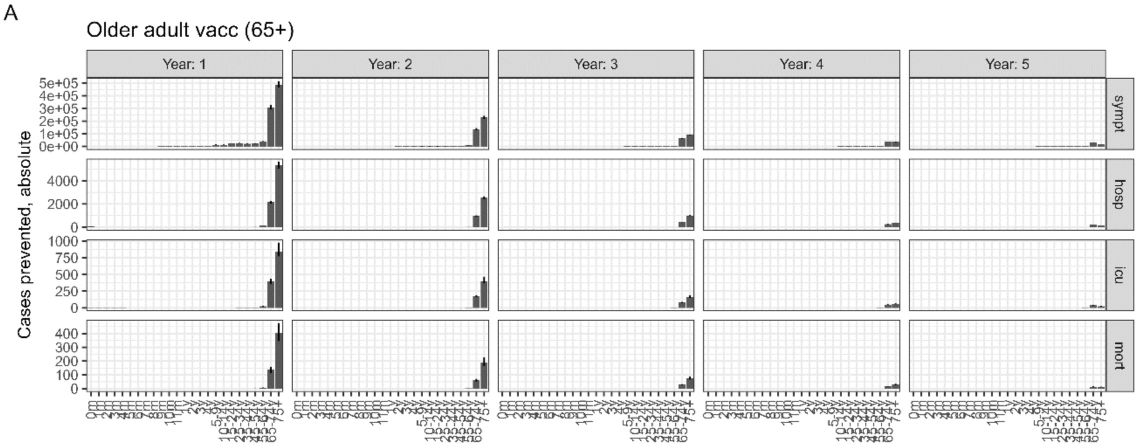

B

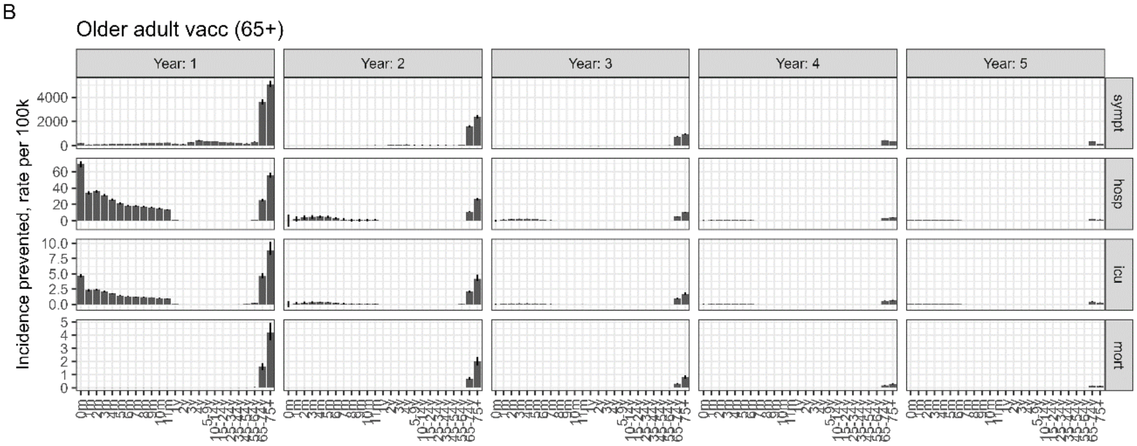

C

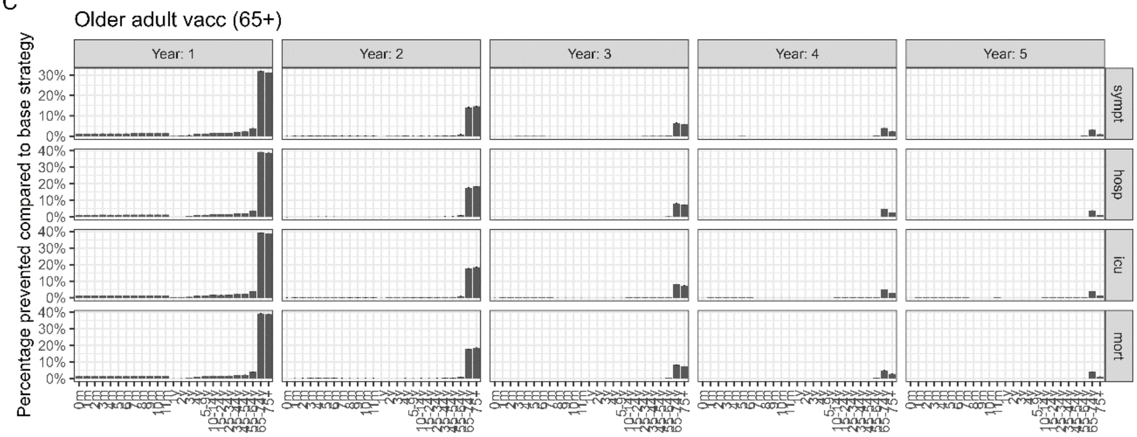

Fig S22: **Model-based prevented incidence of symptomatic RSV and severe disease endpoints per age-group and simulation year for immunisation scenario 5 (older adult vaccination) in 55+ compared to the older adult baseline scenario**. Panel A shows the expected absolute number of prevented cases for each age-group per simulated enpoint (rows) and simulation year (columns). Panel B shows the corresponding rate per 100.000 individuals (i.e., person-years per age-group). Panel C shows the relative reduction in case numbers compared to the baseline scenario.

A

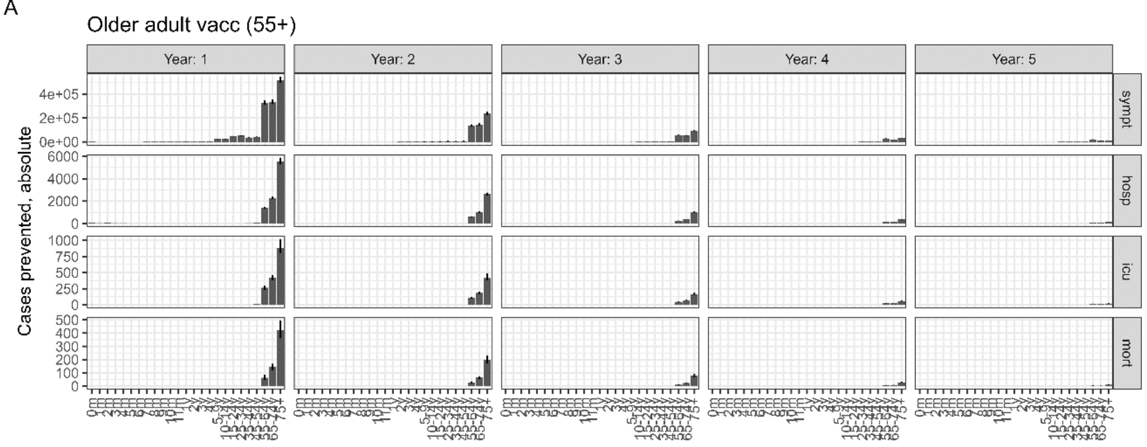

B

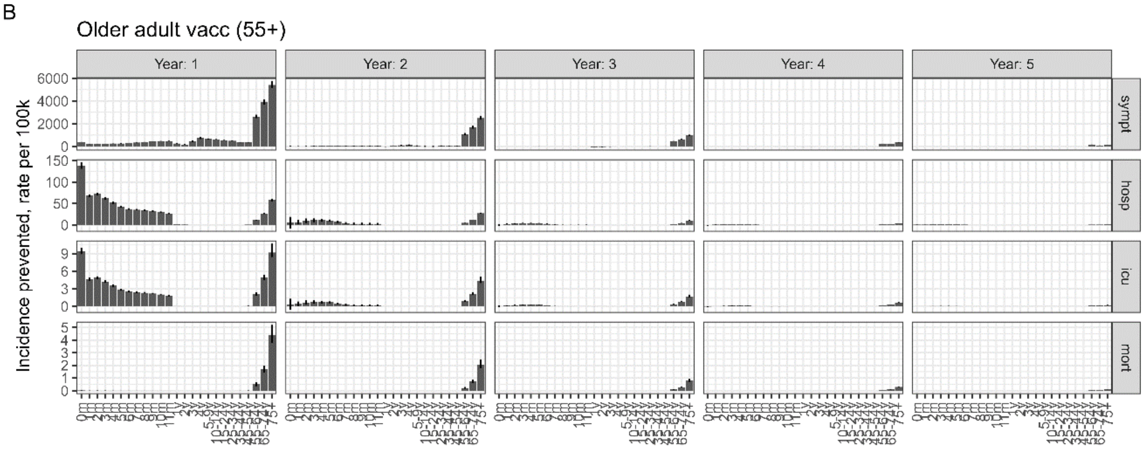

C

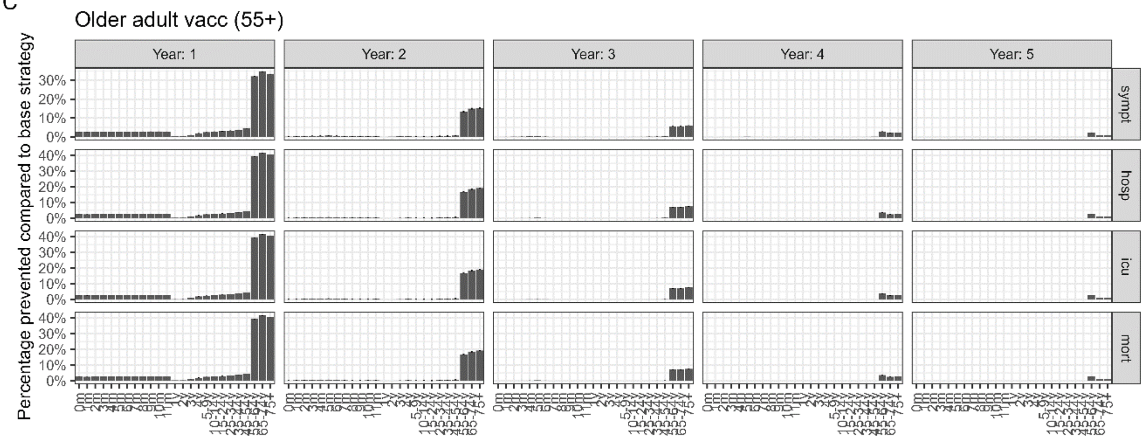

Fig S23: Incremental effect of including additional age-groups into older adult RSV immunisation programme in terms of the prevented total number of symptomatic RSV cases, hospitalisations, ICU-admissions, and deaths in Germany over a 5 years period compared to the current immunisation strategy (Panel A) and the corresponding number needed to vaccinate (Panel B). Shown are the posterior median and associated 95% PI, underreporting of adult RSV-hospitalisations in ICD10-code based RSV-specific hospitalization counts were accounted for based on two different scaling factors: 8 and 14 (purple/green).

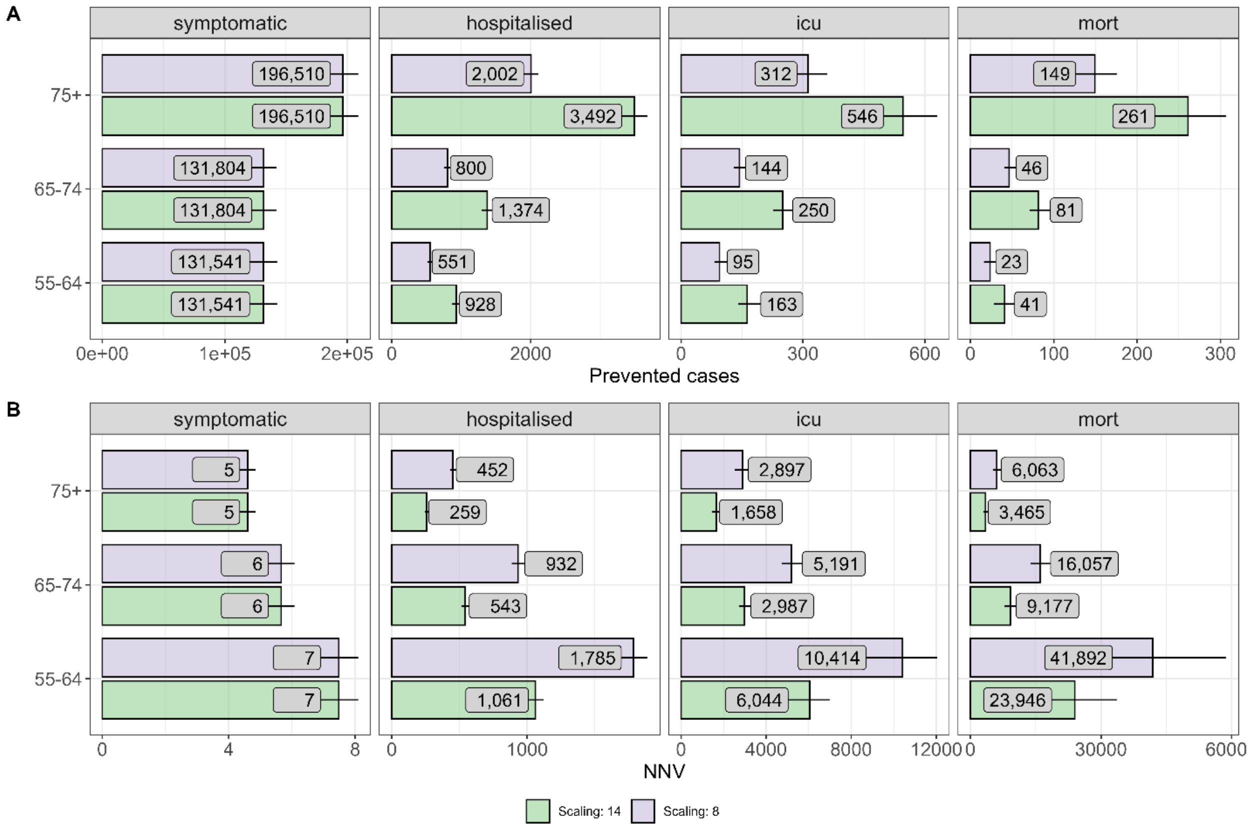

Fig. S24: Forward-simulation results for the one-off older adult RSV vaccination scenario in Germany over five years, with scaling factors of 1-15. Cumulative model results for the older adult RSV immunisation scenarios in terms of the prevented total number hospitalisations, ICU-admissions, and deaths in Germany over a 5 years period compared to the current immunisation strategy (Panel A) and the corresponding number needed to vaccinate (Panel B) based on different scaling factors for adjustment of under-ascertainment in hospitalisation counts for older adults. Shown are the posterior median and associated 95% credible intervals.

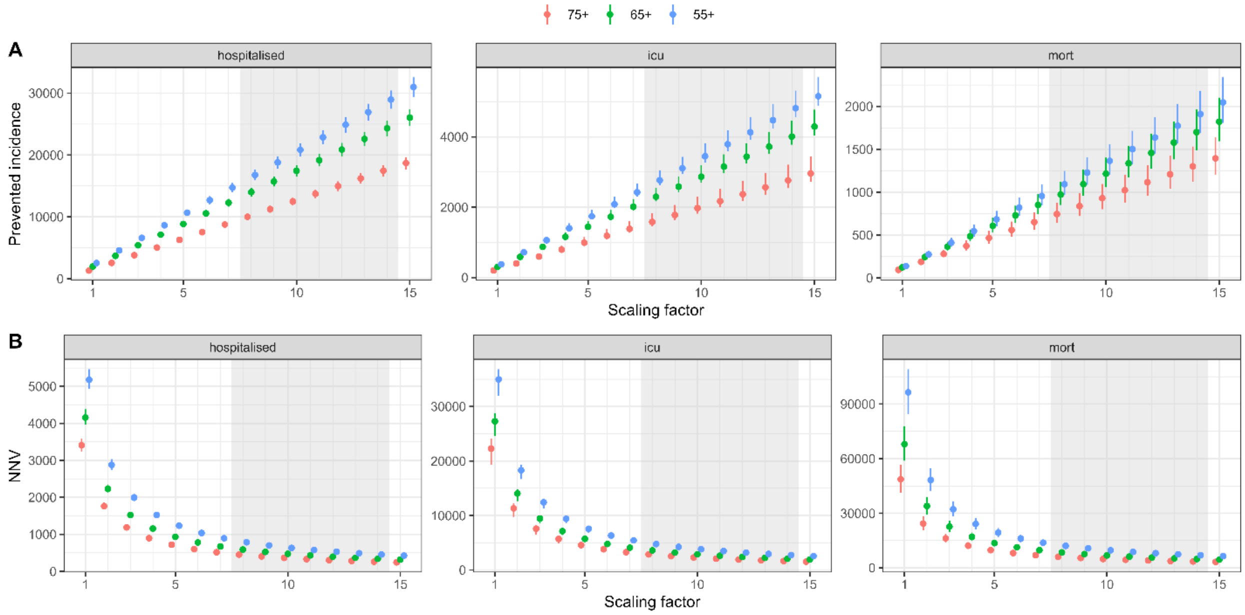
